# Heart rate monitoring using wrist photoplethysmography in Parkinson disease: feasibility and relation with autonomic dysfunction

**DOI:** 10.1101/2025.08.15.25333751

**Authors:** Kars I. Veldkamp, Luc J.W. Evers, Twan van Laarhoven, Yordan P. Raykov, Max A. Little, King Chung Ho, Sooyoon Shin, Lisa Soleymani Lehmann, Bastiaan R. Bloem, Marc A. Brouwer, Jos Thannhauser

## Abstract

**Background:** Cardiac autonomic dysfunction is a prevalent yet underdiagnosed non-motor manifestation in Parkinson disease (PD), and difficult to monitor in daily life. Wrist-worn photoplethysmography (PPG) allows for continuous pulse rate monitoring, with potential to measure cardiac autonomic dysfunction in PD. However, motion artifacts pose significant challenges. We propose a two-step approach to account for PPG motion artifacts, and explore the association between pulse rate characteristics and cardiac autonomic dysfunction in PD.

**Methods:** In 444 people with early PD, we used continuous wrist PPG and accelerometer data, collected during two weeks, with an average wear time of 22h/day. Step 1 filters low-quality segments based on PPG morphology using a logistic regression classifier. Step 2 removes remaining segments with periodic motion artifacts (e.g. introduced by tremor). Weekly aggregates were calculated for resting (day, night) and maximum (day) pulse rates. We studied the effect of filtering on the pulse rate estimates and assessed the relationship between pulse rate aggregates and both subjective and objective autonomic dysfunction measures.

**Results:** Median proportion of high-quality PPG data was 29.2% [IQR: 24.0% to 35.9%] during the day, and 86.1% [IQR: 79.3% to 90.6%] during the night. Proportions of high-quality PPG data were similar across different rest tremor severity and dyskinesia scores. However, filtering out periodic motion artifacts (step 2) reduced overestimation of the maximum HR in persons with mild or severe rest tremor. More symptoms of autonomic dysfunction were associated with a lower maximum pulse rate during the day (β:-0.17, 95% CI: [−0.33, −0.00]), while resting pulse rate showed no association. No significant differences in pulse rate metrics were found between individuals with or without orthostatic hypotension.

**Conclusions:** PPG-based heart rate estimation in PD improves when periodic motion artifacts are accommodated, representing an important step toward developing digital biomarkers of cardiac autonomic dysfunction. Maximum pulse rate in daily life only demonstrated a weak association with autonomic dysfunction, highlighting the need for more specific markers such as pulse rate variability.

**Key contributions/strengths:**

- Careful signal quality assessment, taking into account periodic artifacts by combining PPG and accelerometer
- Careful HR estimation using smoothed-pseudo Wigner-Ville, tested on an independent dataset.
- Evaluation on a unique large PPG dataset from people with PD
- Correlation to the clinical gold standard for autonomic dysfunction
- Pipeline open source available

## Introduction

Non-motor symptoms are an important part of the clinical phenotype of Parkinson disease (PD), and play a crucial role in shaping quality of life among affected individuals[1]. One prevalent yet often underdiagnosed non-motor manifestation of PD is cardiac autonomic dysfunction, including orthostatic hypotension and impaired sympathetic and parasympathetic regulation of heart rate and blood pressure [2–4]. Autonomic regulation of heart rate is affected even in prodromal stages, suggesting early involvement of the autonomic nervous system [5]. Cardiac autonomic failure can be diagnosed through cardiovascular reflex tests, including active standing, tilt-table testing, Valsalva maneuver, and deep breathing [6,7]. These tests are considered as the gold standard for evaluating cardiac autonomic dysfunction, but they are conducted in controlled clinical settings and require specialized equipment and supervision, making them impractical for frequent or long-term monitoring in daily life. Currently, longitudinal assessment of cardiac autonomic symptoms in PD relies primarily on patient-reported questionnaires, which are influenced by the subjective interpretations of both patients and clinicians [8–10]. Therefore, there is need for more objective and continuous methods to assess autonomic symptoms in PD [11–13].

A potential alternative for evaluating autonomic dysfunction in PD involves analyzing heart rate variation. People with PD show lower maximum heart rates, elevated resting heart rates and impaired heart rate recovery after exercise [14,15]. However, current insights into heart rate abnormalities currently stem mainly from episodic heart rate measurements. Wrist photoplethysmography (PPG) can be used to continuous monitor pulse rate, which under typical conditions corresponds to heart rate [16]. This makes PPG an appealing option for daily-life assessment of heart rate patterns as potential digital biomarkers of cardiac autonomic dysfunction in PD.

However, its use in ambulatory settings is technically challenging due to motion artifacts [17], which may be especially relevant in people with PD, who often experience excessive involuntary movements caused by tremor or dyskinesia. Such movements may distort the PPG waveform, reducing the reliability of pulse rate estimation. Therefore, assessing the quality of PPG signals in this population and developing a pipeline to reliably extract pulse rate parameters is a critical first step toward using these methods to monitor autonomic function in daily life.

In this study, our primary objective is to assess PPG signal quality in the PD population by quantifying the prevalence of motion artifacts. We quantify PPG signal quality throughout day and night, focusing on the artifact-inducing effects of motor symptoms on PPG signal quality and corresponding pulse rate estimation. As a secondary, proof-of-principle objective, we explore whether pulse rate parameters derived from daily-life PPG are related to cardiac autonomic dysfunction. To this end, we assess the association of pulse rate parameters with self-reported autonomic symptoms, and compare them between individuals with and without objectively measured orthostatic hypotension.

## Methods

### Study cohort

Data were obtained from the Personalized Parkinson Project (PPP), a single-center cohort study (NCT03364894) including 520 people with early PD (i.e. time since diagnosis ≤5 years) [11]. In brief, participants were monitored in daily life using a wrist-worn sensor device for a minimum of two years (and up to three years in a large subgroup). Participants were also assessed during yearly in-clinic study visits, which included a detailed clinical assessment, MRI, and collection of biosamples.

In the current study, we included participants who completed the baseline clinical assessment and the first two weeks of wearable sensor data collection. The exclusion criteria were insufficient sensor data collection (weekly average wearable wear time < 12 hours per day) and presence of atrial rhythm disorders (as confirmed by screening for cardiac anomalies using electrocardiogram (ECG) recordings (12-lead Holter) or based on medical records). The latter is particularly relevant to ensure that heart rate patterns reflect autonomic function rather than underlying heart rhythm abnormalities.

### Demographics and clinical data

Data on patient demographics, medical history, medication use and symptom severity were obtained from the baseline study visit. To subjectively quantify the severity of autonomic dysfunction, we used the total score on the SCales for Outcomes in PArkinson’s disease - Autonomic Dysfunction (SCOPA-AUT) questionnaire [18]. Objective assessment of cardiac autonomic dysfunction was based on the presence of orthostatic hypotension, as determined during an active standing test [19].

Additionally, the severity of tremor and dyskinesia was obtained from the Movement Disorders Society-Unified Parkinson Disease Rating Scale (MDS-UPDRS) part III and IV assessments, as conducted by trained assessors [20].

### Wearable sensor data

Continuous monitoring of the participants was facilitated using a wearable sensor device (Verily Study Watch, Verily Life Sciences, CA, USA). This watch enables data collection by several sensors, including PPG and accelerometer. During the baseline study visit, participants were instructed on the correct watch placement on the arm, charging, and maintenance. PPG data were obtained at a sampling rate of 30 Hz and accelerometer data at a sampling rate of 100 Hz. In this study, we used the PPG and accelerometer data obtained during the first two weeks of follow-up.

### Wearable sensor data preprocessing

The raw sensor data were stored in the *TSDF* data format to allow for efficient processing [21]. Minor variations in the sampling frequency were resolved by resampling the PPG data through cubic spline interpolation to exactly 30 Hz (PPG) and 100 Hz (accelerometer). Two fourth-order high-pass Butterworth filters with the cut-off frequencies of 0.4 Hz (PPG) and 0.2 Hz (accelerometer) were applied to eliminate offset and trend components in the signals.

### Signal quality assessment

We employ an approach to filter out PPG segments with significant motion artifacts which could impede reliable pulse rate estimation using the combined outputs of the following steps:

1. Assessment of PPG morphology
2. Periodic motion artifact removal

In the first step, segments are filtered out based on the typical PPG morphology. In the second step, remaining segments with periodic motion artifacts (e.g. introduced by tremor), which mimic pulse waves, were filtered out by combining the PPG and accelerometer data. The schematic representation of this approach is depicted in Figure 1.

**Figure 1:**
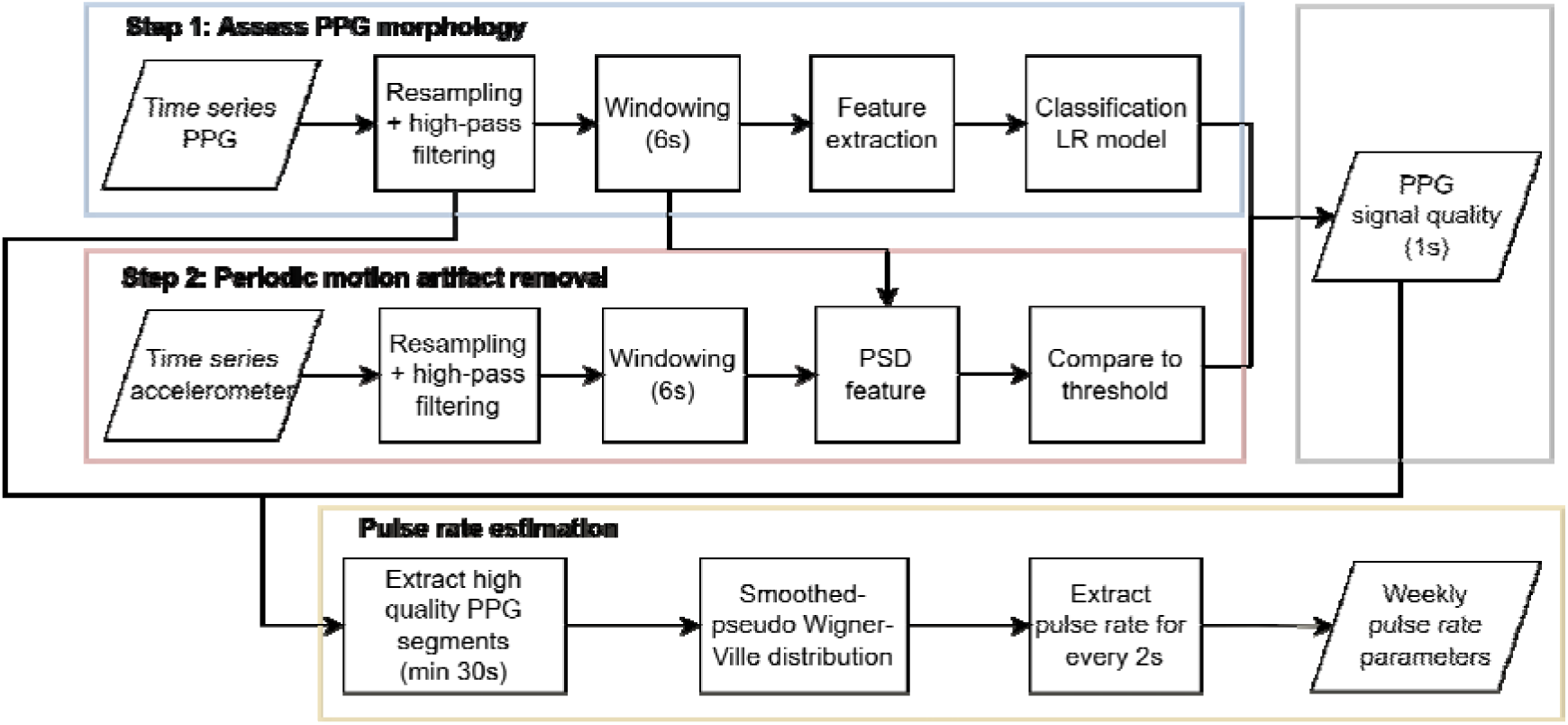
An overview of the signal processing pipeline for PPG signal quality and pulse rate estimation. The pipeline consists of a PPG signal quality filter and a pulse rate estimation method: 1) signal quality filter, combining PPG morphology classification (step 1) and periodic motion artifact detection (step 2) using accelerometer data to obtain per-second quality labels; and 2) pulse rate estimation from high-quality PPG segments using the smoothed pseudo Wigner-Ville distribution. Pulse rate is estimated for every 2s and summarized weekly. LR = logistic regression, PPG = photoplethysmography; PSD = power spectral density.

### Step 1: Assessment of PPG morphology

We assessed PPG morphology based on the typical sinusoidal pulse waves with discernible peaks from which pulse rate can be determined [22–24]. To detect typical PPG morphology, we employed a machine learning modelling approach – a logistic regression (LR) classifier. The LR classifier was trained using an annotated dataset consisting of PPG data from 20 PPP subjects (training set, not used in the other evaluations), selected through stratified sampling based on age, sex, and rest tremor scores. The PPG signals were annotated by visual inspection by two experienced investigators (KIV, JT) following a standardized annotation protocol, which included a detailed definition for the PPG morphology of low-versus high-quality segments (Supplementary Methods 1.1.1.2). First, the two investigators independently annotated the same segments and discussed any disagreements to refine the annotation protocol. After sufficient agreement was reached (kappa of 0.8), the remaining segments were annotated by a single investigator. A detailed description of the annotation process can be found in Supplementary Methods 1.1.1.

The LR model to classify PPG morphology was trained using window-based time and frequency features. L1 regularization was applied for feature selection to enhance classifier efficiency. The performance was evaluated using leave-one-subject-out cross-validation. Compared to the visual annotations, the classifier’s accuracy was 98.4% (SD: 0.95%), with a sensitivity of 98.2% (SD: 1.32%) and a specificity of 98.5% (SD: 1.32%).

A detailed description of the training process, including feature selection and the specific features used in the LR classifier, can be found in Supplementary Methods 1.1.2-1.1.4.

### Step 2: Periodic motion artifact removal

Due to the sensitivity of PPG to motion artifacts, we hypothesized that periodic movements such as tremor can cause periodic artifacts very similar to the typical pulse wave in the PPG signal. During the annotation process, we indeed found several examples of oscillations in the PPG signal that were accompanied by oscillations of the same frequency in the accelerometer signal (see Figure 2 for one example). These examples occurred in the low rest tremor range (around 2.5 to 3 Hz), which is still within the physiological pulse rate range.

**Figure 2:**
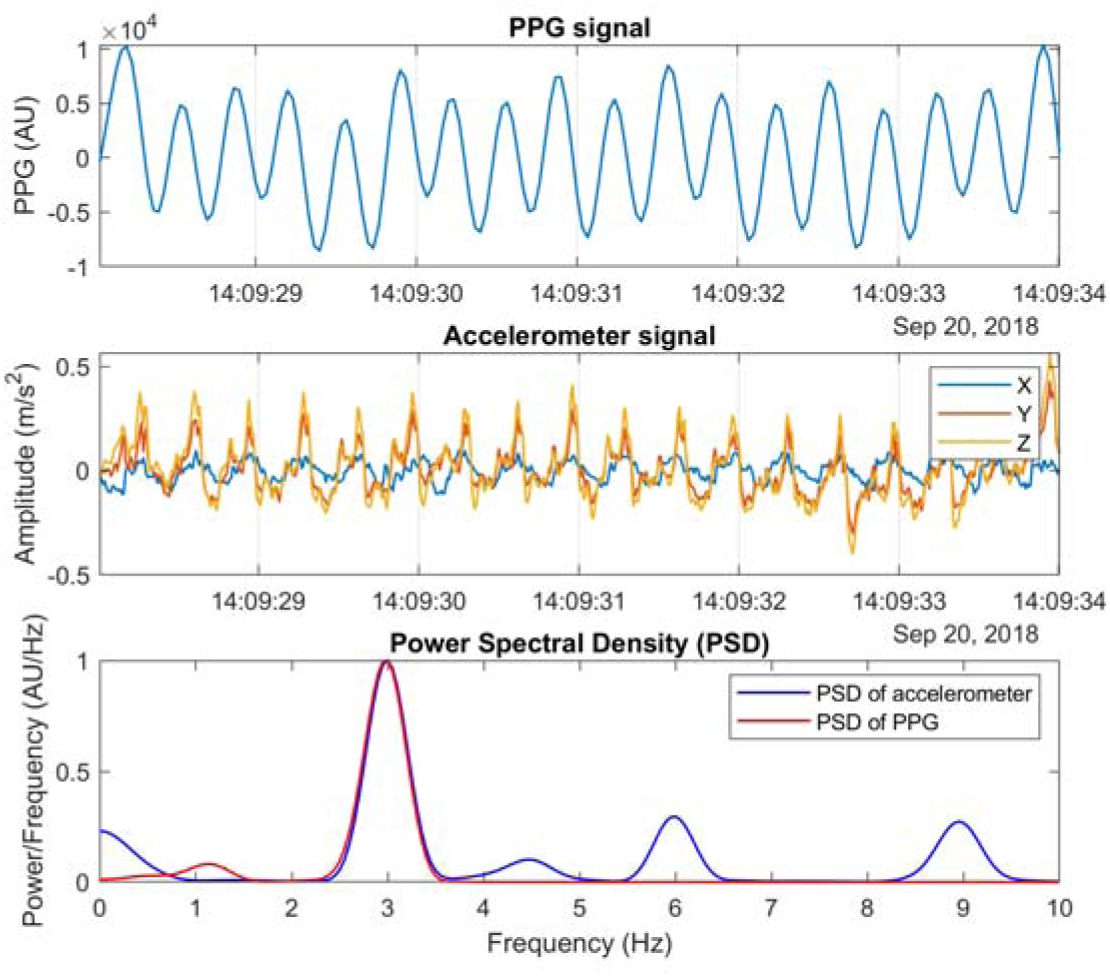
Possible leakage of periodic movements into the PPG signal. Upper panel: PPG signal, middle panel: three axes of the accelerometer, lower panel: power spectral density of PPG and the sum of the three accelerometer axes. The dominant frequency in both signals is +/- 3 Hz, which falls within both the physiological pulse rate range and

To filter out PPG segments with such periodic artifacts, we combined the PPG and accelerometer signals. We considered a segment to be a periodic artifact if the dominant frequency of the PPG matched the dominant frequency of the accelerometer. To identify these artifacts, we calculated the window-based relative power in the accelerometer at the dominant PPG frequency (±0.05 Hz) and its first harmonic (±0.05 Hz). If the relative power exceeded a threshold of 0.10 the window was classified as an artifact. The threshold was selected based on the training dataset. Supplementary Methods 1.2 provide more details on the calculation of the relative power and the threshold selection.

### Composite signal quality classification

Both steps were applied to 6s windows with a 5s overlap, after which the final signal quality classification was made for every 1s. A window was classified as high-quality, only if both steps classified it as such. To produce the final quality prediction for every 1s, majority voting was applied to the overlapping windows. More details can be found in Supplementary Methods 1.3.

### Impact of tremor and dyskinesia on signal quality

To investigate the impact of tremor and dyskinesia on PPG signal quality, participants were stratified into sub-groups based on their rest tremor scores on the device-sided arm (item 3.17) and combined dyskinesia scores (items 4.1 and 4.2) from the MDS-UPDRS at the baseline visit. Rest tremor scores were categorized into three groups: no tremor (score of 0), mild tremor (score of 1), and severe tremor (score of 2 or higher). Similarly, dyskinesia scores, derived from both the duration and functional impact of dyskinesias, were divided into three groups: no dyskinesia (combined score of 0), mild dyskinesia (combined score of 1), and severe dyskinesia (combined score of 2 or higher). Signal quality estimates were analyzed separately for daytime hours (08:00 – 21:59) and nighttime hours (00:00 – 05:59).

### Pulse rate estimation

Following evaluation of the PPG signal quality, we proceeded with pulse rate estimation, as depicted in Figure 1. For this, 30 consecutive high-quality 1s windows were used, as determined by the signal quality algorithm. To identify the most suitable method for pulse rate estimation from PPG, we compared various approaches using the PPG DaLiA dataset[25], with ECG as reference. The highest accuracy was obtained using smoothed-pseudo Wigner-Ville Distribution (SPWVD) time-frequency analysis, hence we used the SPWVD for all subsequent pulse rate estimates. See Supplementary Methods 1.4.1-1.4.6 for the results of the comparison.

Using the SPWVD method, we estimated the pulse rate for every 2s. An example of this approach is provided in Supplementary Methods 1.4.7. Pulse rate estimates were divided into either daytime (08:00 – 21:59) or nighttime (00:00 – 05:59) and aggregated per week into three parameters: resting pulse rate (day and night) and maximum pulse rate during the day. The resting pulse rate was defined as the most frequently occurring pulse rate value across high-quality windows within the respective time windows. To ensure a meaningful mode calculation, pulse rate estimates were assigned to bins of 1 beat per minute, based on the approximate frequency resolution from the 30-second SPWVD window. Maximum pulse rate during the day was defined as the 99% percentile of all pulse rate estimates.

### Impact of periodic artifact removal on pulse rate estimates

To evaluate the impact of periodic motion artifacts on pulse rate estimations, we compared the results of two configurations of the processing pipeline: (1) using only PPG morphology for high-quality segment selection (step 1), and (2) using the combined PPG and accelerometer approach to also account for periodic artifacts (steps 1 and 2). The two seconds pulse rate estimates and weekly pulse rate aggregates derived from both configurations were analyzed across the previously defined study groups for rest tremor and dyskinesia severity.

### Effect of autonomic dysfunction on pulse rate parameters

We used a linear regression model to investigate the effect of autonomic symptom severity, as assessed both subjectively by the continuous sum score of the SCOPA-AUT questionnaire and objectively based on the presence of orthostatic hypotension, on the weekly pulse rate parameters. The pulse rate parameters served as dependent variables in the regression analyses. To reduce bias and strengthen the interpretation of this effect estimate in an observational setting [26], we constructed a direct acyclic graphs (DAG) to identify potential confounding factors known to influence both pulse rate parameters and autonomic dysfunction (see Supplementary Methods 1.5). Based on this DAG, we adjusted the regression models for age, sex, baseline physical activity (assessed by the Physical Activity Scale for the Elderly; PASE), and medication (specifically betablockers). Statistical significance for comparisons between study groups (e.g. tremor and dyskinesia severity) was assessed using one-way ANOVA or Kruskal-Wallis tests, whichever was appropriate. For within-subject comparisons (e.g., pulse rate parameters before and after periodic artifact removal), paired t-tests or Wilcoxon signed-rank tests were applied, whichever was appropriate. A significance threshold of p < 0.05 was used. The linear regression models were implemented using the LM function in R (RStudio, version 4.3.2).

## Data availability

Data from the Personalized Parkinson Project used in the present study were retrieved from the PEP database (https://pep.cs.ru.nl/index.html). The PPP data is available upon request via. More details on the procedure can be found on the website www.personalizedparkinsonproject.com/home.

## Code availability

Code for the running the individual PPG signal processing pipeline (preprocessing, signal quality assessment and pulse rate estimation) is available in the ParaDigMa toolbox: https://doi.org/10.5281/zenodo.15223364. The code to generate the aggregated results in this study is publicly available in the Git repository: https://github.com/biomarkersParkinson/PPP_PPG_feasibility.git.

## Results

### Study population

The baseline characteristics of the study population are presented in Table 1. From the 520 participants in the PPP, 34 participants had insufficient data recordings in week 0 (n=10), week 1 (n=17) or both (n=7). Moreover, 22 participants had proven rhythm disorders on the screening ECG. 20 participants selected using stratified sampling were used for training the signal quality algorithm. The remaining 444 participants were included in the main analyses.

**Table 1:** Baseline characteristics of the study population. Data are means (SD) or numbers (%)

|  | N = 444 |
| --- | --- |
| Age (years) | 61.4 (9.0) |
| Sex (number of women) | 187 (42.1%) |
| Time since diagnosis (months) | 31.4 (17.5) |
| SCOPA-AUT score, range 0-69 | 14.5 (7.2) |
| **Orthostatic hypotension** | 68 (15.3%) |
| **MDS-UPDRS Part III OFF score, range 0-132** | 33.2 (12.9) |
| **Use of beta blockers** | 40 (9.0%) |
| **PASE score, range 0-400** | 173.2 (81.2) |
| **Resting pulse rate – day (min<sup>-1</sup>)** | 66.5 (9.2) |
| **Resting pulse rate – night (min<sup>-1</sup>)** | 61.4 (8.0) |
| **Maximum pulse rate (min<sup>-1</sup>)** | 87.1 (13.0) |
| **Daily sensor data availability week 0 (h/day)** | 22.0 (1.1) |
| **Daily sensor data availability week 1 (h/day)** | 22.1 (1.2) |

Participants had a mean age of 61.4 years (standard deviation, SD: 9.0), of whom 187 (42.1%) were women. Mean time since PD diagnosis was 31.4 (SD: 17.5) months. Orthostatic hypotension, as measured by the active standing test, was present in 68 participants. MDS-UPDRS Part III scores (in the off state) reflected mild to moderate disease severity (33.2, SD: 12.9). The synchronized sensor data (accelerometer + PPG) available for analysis comprised 22.0 hours per day per participant (SD: 1.1) in week 0 and 22.1 hours per day (SD: 1.2) in week 1.

### Signal quality - descriptives

Signal quality was defined using a two-step algorithm combining PPG signal morphology and detection of periodic motion artifacts. This approach is visually summarized in Figure 1, providing a clear overview of the criteria used to identify high-quality segments.

In Figure 3, the median (IQR) proportion of high quality PPG signals is shown across every hour of the circadian cycle. During daytime hours, the median proportion was 29.2% [24.0%, 35.9%]; while during nighttime hours, it was 86.1% [79.3%, 90.6%]. We observed a similar pattern in the second study week (Supplementary Figure 5, Supplementary Table 3). Removing periodic artifacts resulted in an overall reduction of 0.10% of the median proportion of high-quality PPG signals during the day (Supplementary Figure 6, Supplementary Table 3).

**Figure 3:**
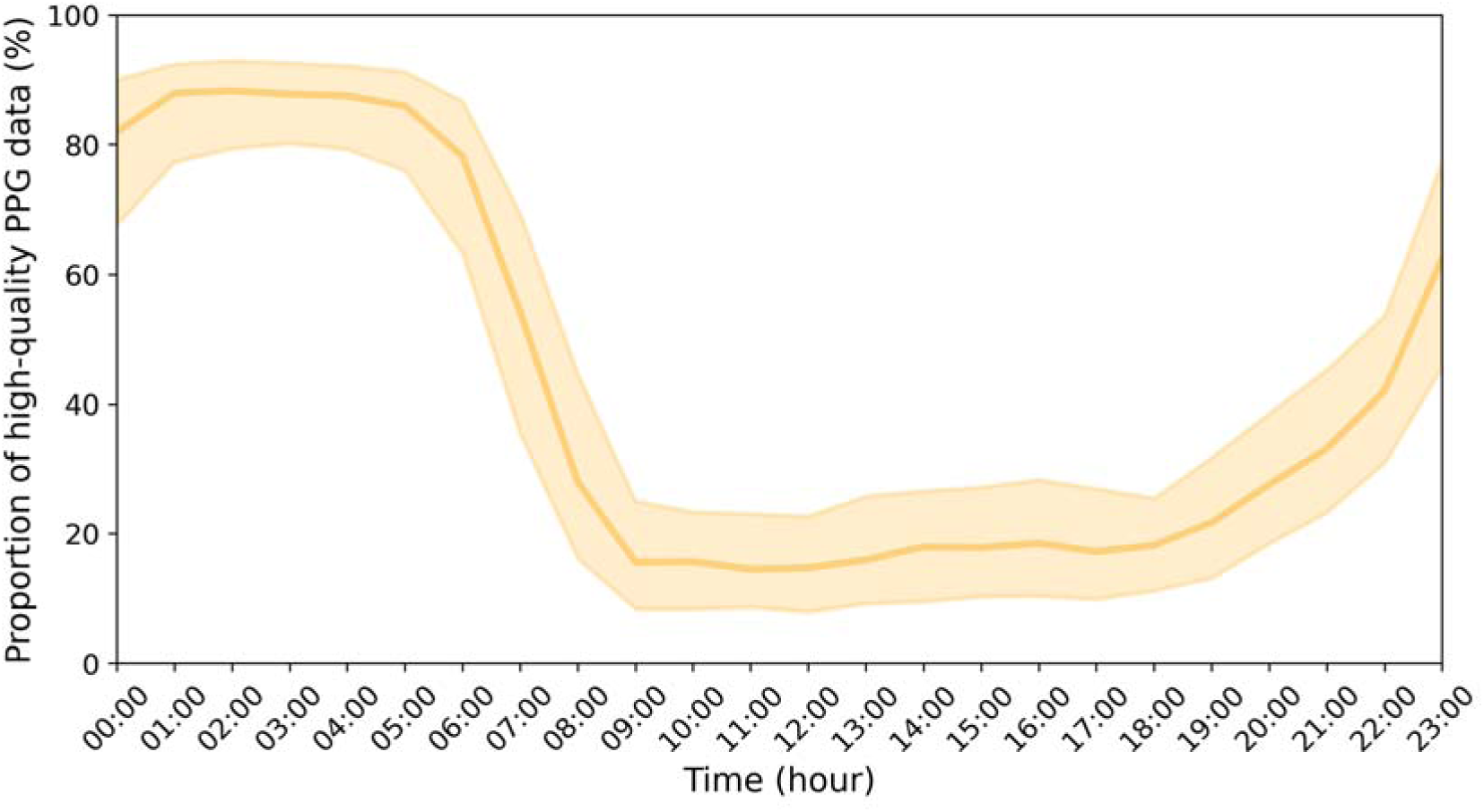
Proportion of high-quality PPG data across the circadian cycle in the first study week. Data are represented as medians + IQR.

### Impact of tremor and dyskinesia on signal quality

No significant differences in day-time data quality were found among the no tremor, mild tremor, and severe tremor groups [*F*(2,441)=0.19, *p*=0.83; Figure 4]. Similarly, no significant differences in day-time data quality were found among the no dyskinesia, mild dyskinesia and severe dyskinesia groups [*F*(2,441)=1.07, *p*=0.34; Figure 5]. All comparisons between study groups based on tremor and dyskinesia can be found in Supplementary Tables 3 and 4.

**Figure 4:**
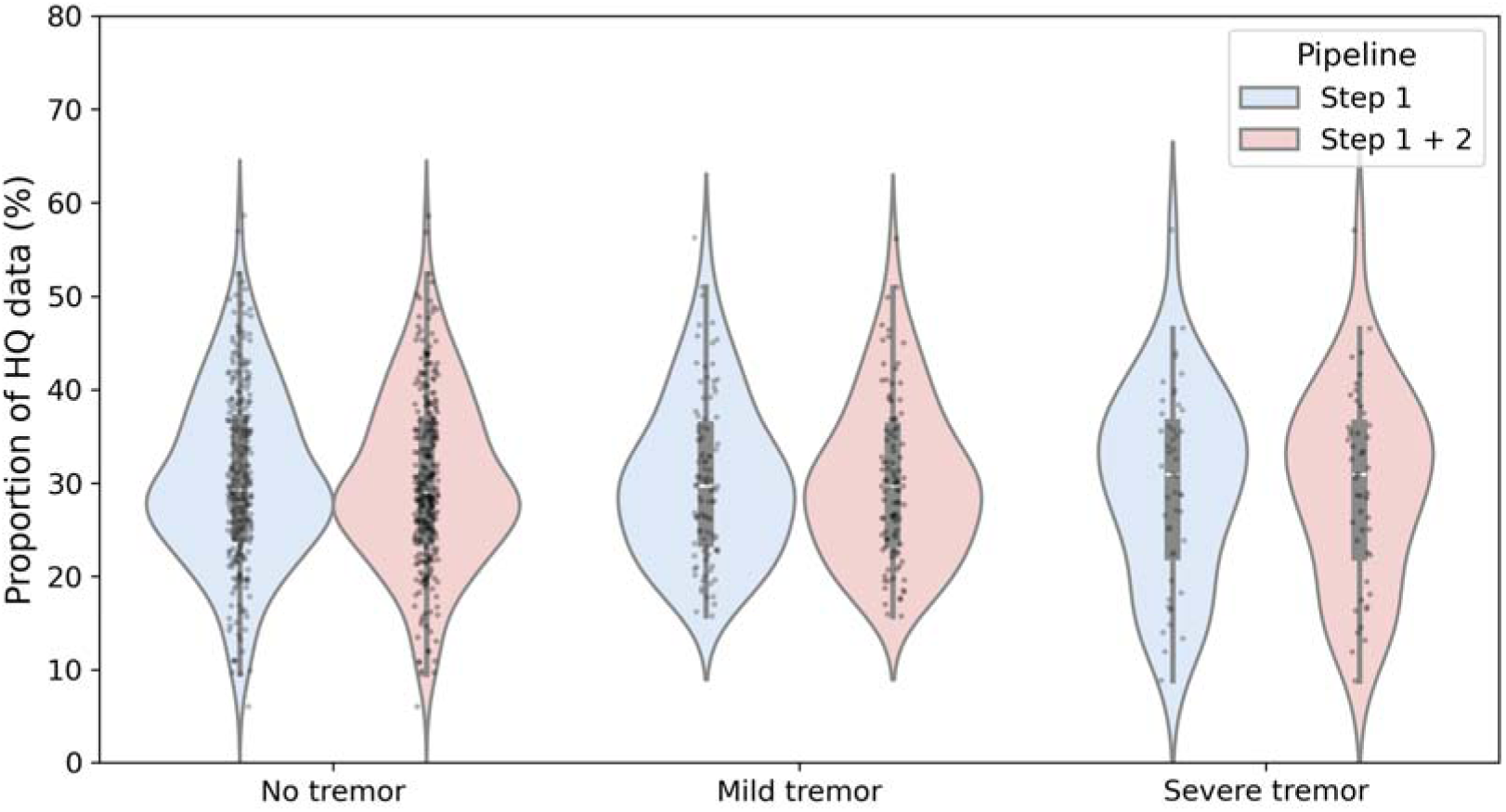
The proportion of high-quality PPG data in relation to tremor severity (MDS-UPDRS 3.17) in the first study week. The different colors represent the data quality when assessing solely PPG morphology (blue) or also incorporating periodic artifact removal using the accelerometer (red).

**Figure 5:**
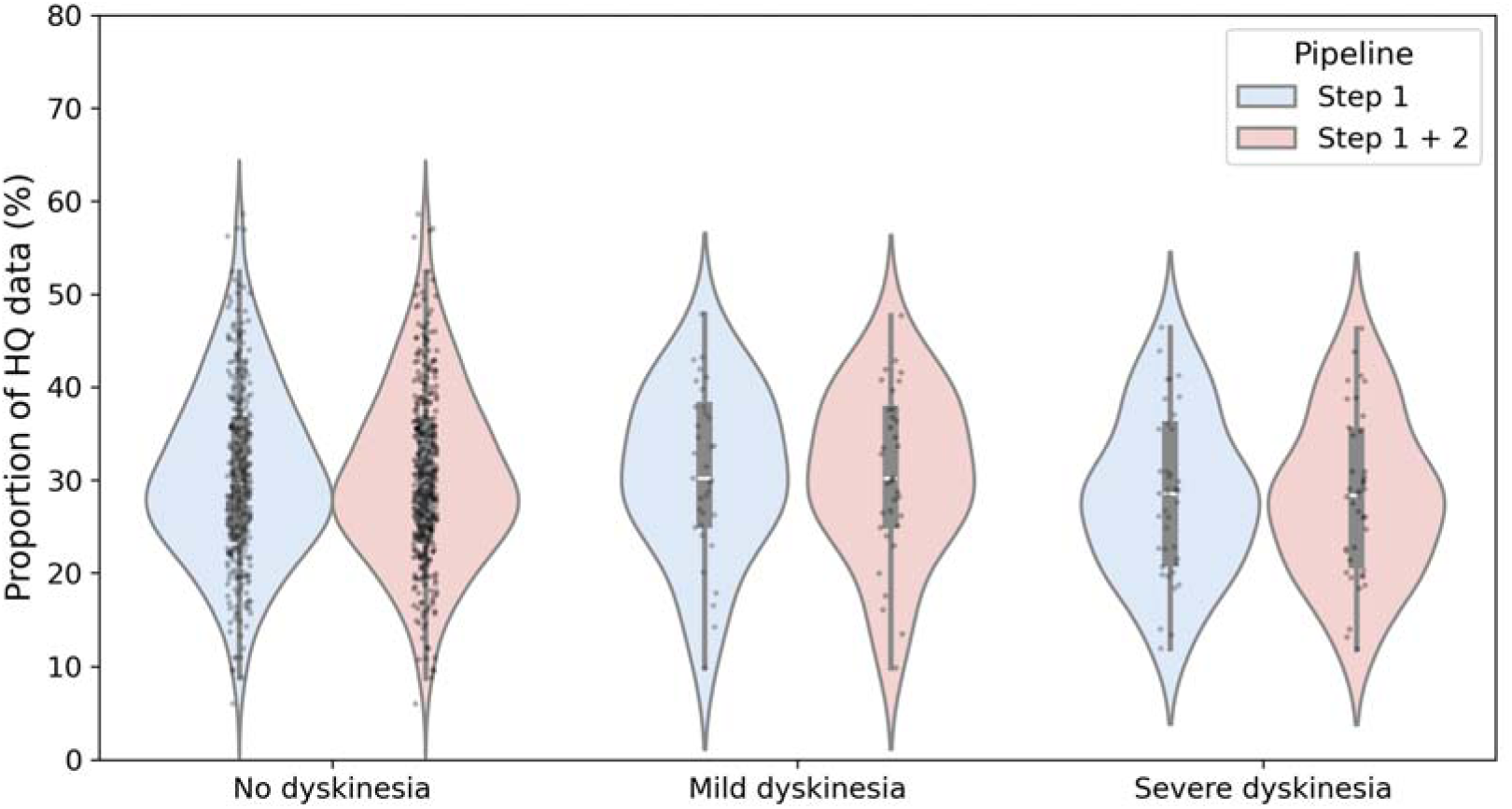
The proportion of high-quality PPG data in relation to dyskinesia severity (MDS-UPDRS 4.1 + 4.2) in the first study week. The different colors represent the data quality when assessing solely PPG morphology (blue) or also incorporating periodic artifact removal using the accelerometer (red).

### Impact of periodic artifact removal on pulse rate estimates

Next, we assessed the effect of periodic artifact removal on two-second pulse rate estimates in the physiological range (40-180 beats per minute). Pulse rate was estimated every two seconds using smoothed pseudo Wigner-Ville distribution (SPWVD) time-frequency analysis, applied to 30-second segments of high-quality PPG signals.

Figure 6 illustrates the mean absolute removal of pulse rate estimates per subject across the different tremor groups in the first study week. We identified and removed more periodic motion artifacts in individuals with higher tremor scores, particularly in the >160 bpm range. No difference is seen between the different dyskinesia groups (Supplementary Figure 7). Furthermore, in all study groups, low-frequency (40-120 bpm) periodic motion artifacts were removed, with the highest proportion in the 40-80 bpm range.

**Figure 6:**
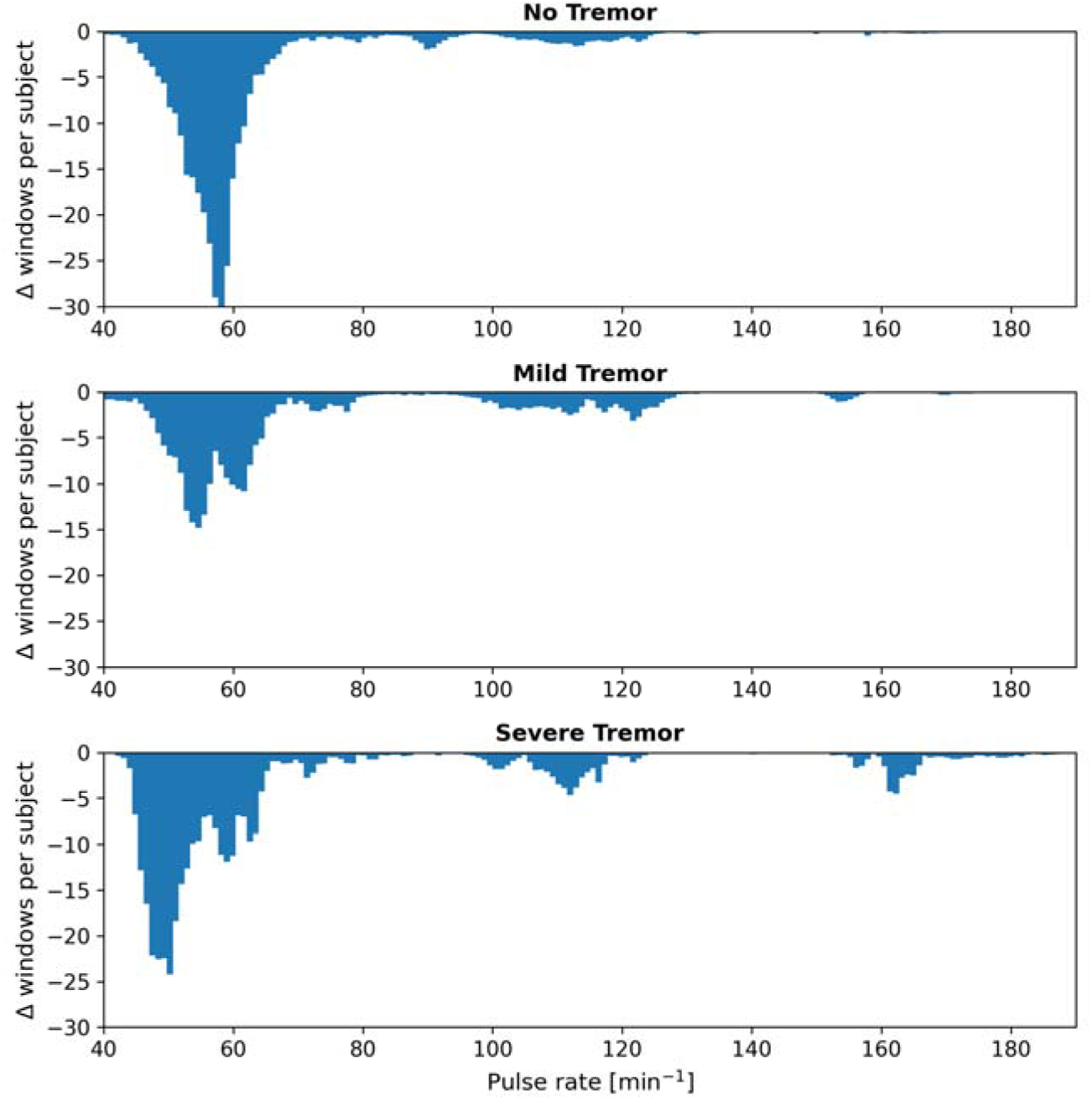
Removed pulse rate estimates by applying step 2 of the signal quality algorithm, for the different tremor groups. The data show the mean change in counts per subject for each pulse rate value. People with a higher tremor score show more exclusion of higher pulse rate estimates.

Figure 7 shows a representative example of a data fragment from a severe tremor subject where periodic movement artifacts influenced the PPG signal. By applying the periodic motion artifact filter in step 2 of the signal quality algorithm, this segment is excluded from further pulse rate analysis.

**Figure 7:**
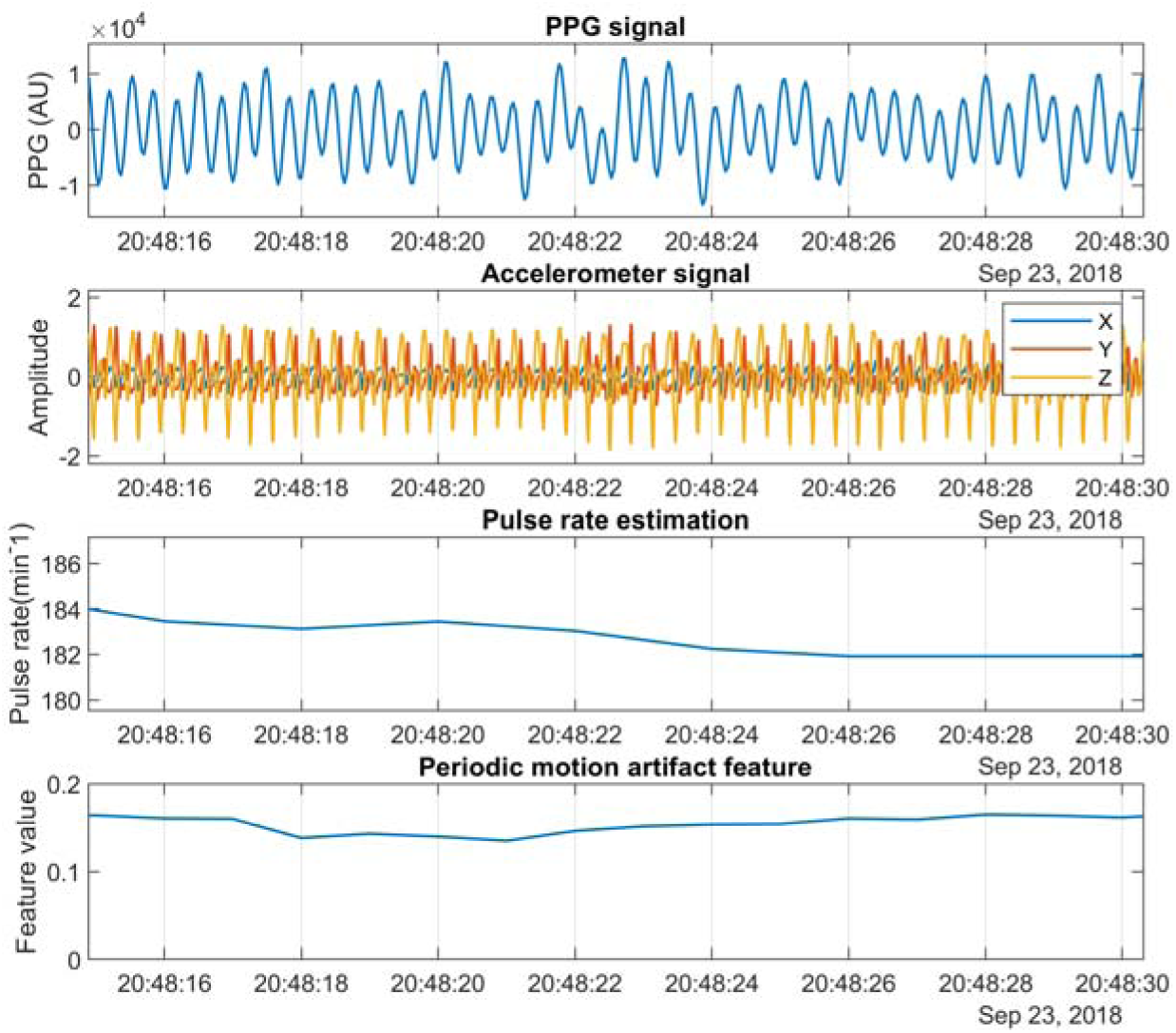
Data fragment of periodic artifact leaking into PPG in a severe tremor subject. In the upper two panels, the PPG signal shows substantial alignment with acceleration, indicating that a potential motion artifact is present in the PPG signal. Without artifact removal, the extracted pulse rate of the PPG signal is approximately 180 beats per minute (third panel). However, the relative power in the accelerometer at the dominant PPG frequency (fourth panel) exceeds the motion artifact threshold of 0.10, leading to the exclusion of this estimate from further analysis.

Next, we assessed the impact of periodic artifact removal on the weekly aggregated pulse rate parameters. After filtering, the mean resting pulse rate was 61.4 min^-1^ (SD: 8.0) at night and 66.5 min^-1^ (SD: 9.2) during the day. The mean maximum pulse rate was 87.1 min^-1^ (SD: 13.0). Full distributions of all pulse rate parameters are shown in Supplementary Figure 8. The effect of periodic motion artifact removal (Step 2) on the aggregated maximum pulse rate across the three tremor groups is illustrated in Figure 8. There were significant differences in maximum pulse rate between the two configurations across all tremor groups. In participants without tremor, the mean maximum pulse rate decreased from 87.2 bpm to 85.2 bpm after periodic motion artifact removal, a decrease of 2.0 bpm (SD 8.2; p < 0.001). In those with mild tremor, the mean maximum pulse rates dropped from 91.3 bpm to 87.5 bpm after, a difference of 3.8 bpm (SD 11.3; p = 0.001). Participants with severe tremor showed a decrease from 97.6 bpm before and 92.7 bpm, representing a difference of 4.9 bpm (SD 14.7; p = 0.02). On an individual level, several maximum pulse rates in all groups were affected by more than 30 BPM. We saw a similar pattern for the three different dyskinesia groups (Supplementary Figure 10).

**Figure 8:**
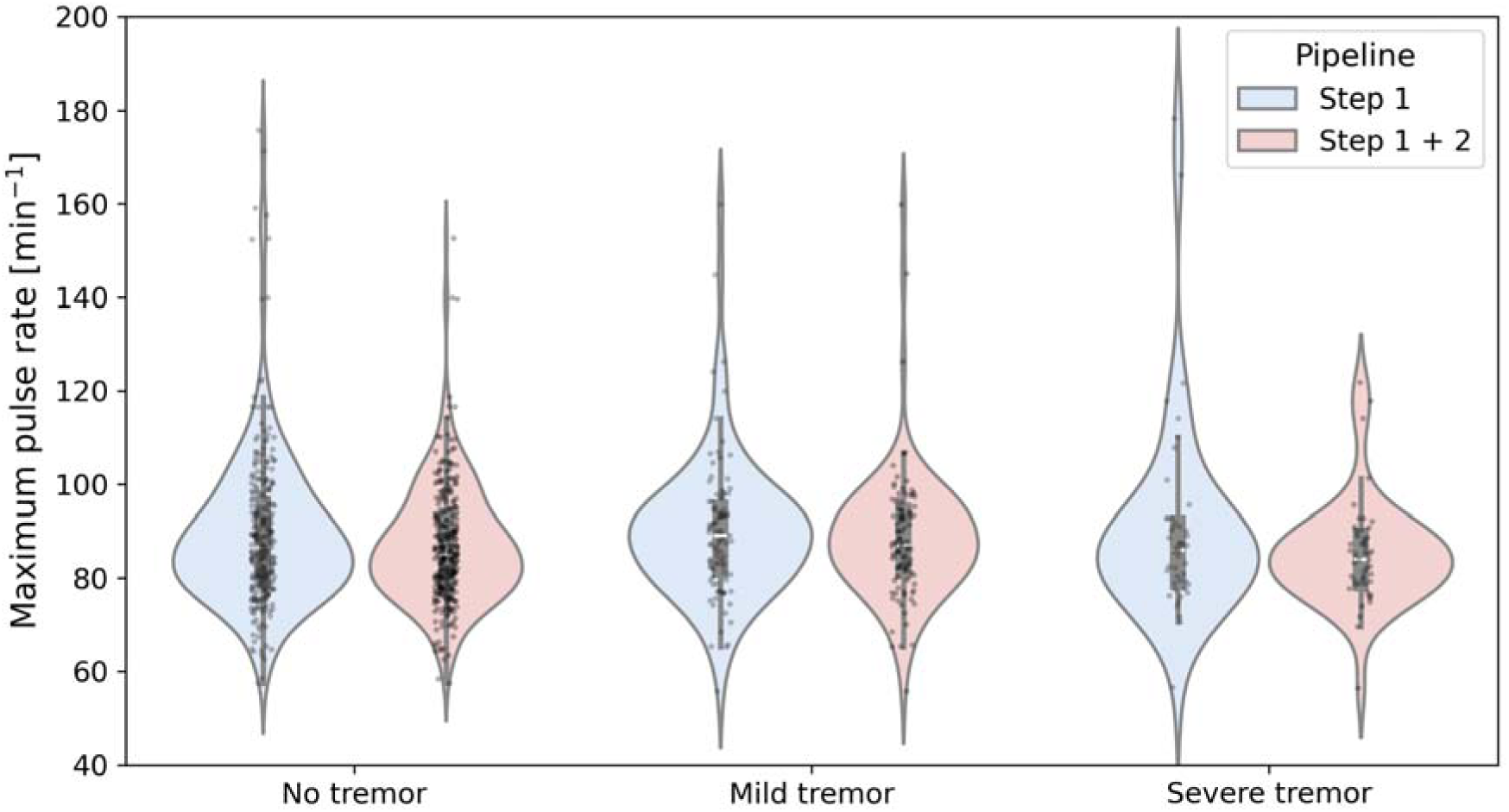
The maximum pulse rate in relation to tremor severity (MDS-UPDRS 3.17) in the first study week. The different colors represent the maximum pulse rate when assessing solely PPG morphology (blue) or also incorporating periodic artifact removal using the accelerometer (red). While filtering for periodic motion artifacts significantly affects the aggregated maximum pulse rate at the group level, its effect was particularly substantial at a individual level – especially in the severe tremor group.

In contrast to the maximum pulse rate, individual resting pulse rate parameters were not affected by removal of periodic motion artifacts across tremor and dyskinesia groups (Supplementary Figure 11-14).

### Effect of autonomic dysfunction on pulse rate parameters

Individuals with more (severe) symptoms of autonomic dysfunction had a lower maximum pulse rate during the day in both the first study week (β:-0.17, 95% CI: [−0.33, −0.00]; Figure 9) and the second study week (β:-0.26, 95% CI: [−0.41, −0.04]; Supplementary Figure 15). In contrast, autonomic symptom severity was not associated with resting pulse rate during either the night (β:0.02, 95% CI: [−0.08, 0.13]) or the day (β:-0.02, 95% CI: [−0.13, 0.10]) in the first study week (Figure 9). Similar results were obtained in the second study week (Supplementary Figure 15).

**Figure 9.**
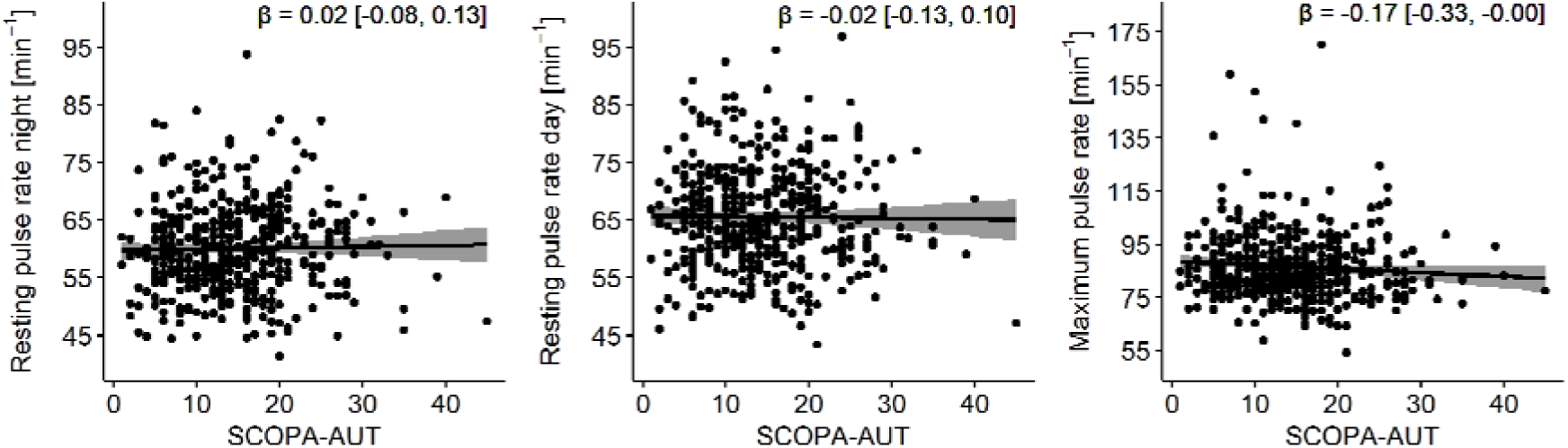
Multivariate regressions between the pulse rate parameters in the first study week and SCOPA-AUT. Every data point is corrected using the following covariates: age, sex, use of betablockers and baseline physical activity. SCOPA-AUT = SCales for Outcomes in PArkinson’s disease - Autonomic dysfunction.

Orthostatic hypotension had no significant effect on the maximum pulse rate in the first study week (β:-2.71, 95% CI: [−6.18, 0.76]) and the second study week (β:-2.56, 95% CI: [−6.43, 1.30]; Supplementary Figure 16). Furthermore, orthostatic hypotension had no effect on the resting pulse rate during the night (β:-0.84, 95% CI: [−2.96, 1.29]) or the day (β:-1.98, 95% CI: [−4.41, 0.44]). Similar results were obtained in the second study week (Supplementary Figure 16).

**Figure 10.**
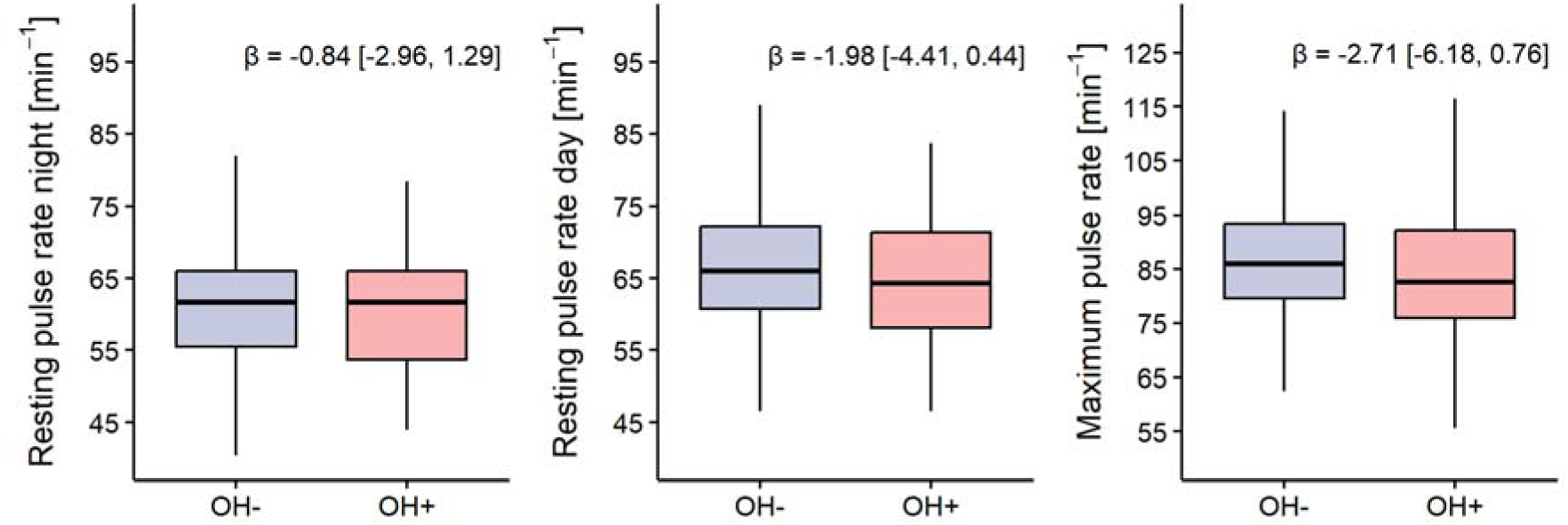
Multivariate regressions between the pulse rate parameters in the first study week and the presence of orthostatic hypotension. Pulse rate parameters are corrected using the following covariates: age, sex, use of betablockers and baseline physical activity. OH = orthostatic hypotension.

## Discussion

This study explored the feasibility of wrist-worn PPG for measuring pulse rate parameters in people with PD during daily life and to assess the effect of autonomic dysfunction on these parameters. First, we addressed the problem of PPG motion artifacts in the PD population. Our findings indicate that the severity of tremor and dyskinesia does not affect the overall availability of high-quality PPG data. However, periodic motion artifacts, such as those caused by tremors, can impact pulse rate estimates. Segments with such periodic artifacts can be filtered out by combining PPG and accelerometer signals, reducing overestimation of the maximum pulse rate at the individual level with minimal data removal. Second, we studied the association between autonomic dysfunction on pulse rate parameters. We found a weak association of severity of autonomic symptoms with weekly maximum pulse rate, but no effect on weekly resting pulse rate. We did not find an effect of orthostatic hypotension on the pulse rate parameters, although the maximum pulse rate was numerically lower in the orthostatic hypotension group. These findings indicate that wrist-worn PPG has the potential to capture features related to cardiac autonomic dysfunction in people with PD. However, further research is needed to identify more specific and sensitive pulse rate (variability) markers to monitor cardiac autonomic dysfunction.

Compared with existing literature, our study provides novel insights into the impact of periodic motion artifacts on the quality of PPG signals in people with PD. The impact of these artifacts on daily life assessments had not been studied before. We demonstrate that filtering out such artifacts (caused by e.g. tremor) minimally reduces the proportion of high-quality PPG data, but it can improve pulse rate estimation in segments affected by periodic motion artifacts. The extraction of pulse rates from PPG signals can be approached using traditional signal analysis methods [22,23,27] or more advanced techniques, such as deep learning combined with sensor fusion (accelerometer and PPG)[28]. Traditional methods, as employed in this study, typically focus on identifying and extracting high-quality PPG signals prior to pulse rate estimation[24]. However, most studies focus solely on PPG signal characteristics, whereas our study also integrates accelerometer data to explicitly account for periodic motion artifacts. This approach is particularly important in the context of PD, as our results suggest that tremor-related motion can mimic physiological patterns in the PPG signal. This insight may extend to broader contexts, such as lower-frequency periodic movements during e.g. gait (around 1 Hz) [29]. This is supported by our data, showing that most removed periodic artifacts, regardless of tremor group, were concentrated in the 40-80 bpm range (0.7-1.3 Hz which corresponds to gait). Although sophisticated approaches like statistical models and machine learning, including deep learning incorporating frequency characteristics of both PPG and accelerometer, have been explored primarily in sports research, they have not yet been specifically developed for diseased populations [28,30,31]. We posit that such approaches are suited to also address PD-specific challenges. Future research should explore these methods, as they may enhance pulse rate estimation reliability during lower-quality PPG segments encountered in daily activities.

Previous lab-based studies demonstrated that people with PD have a lower maximum heart rate during exercise compared to controls due to autonomic dysfunction. We extended this by examining the effect of cardiac autonomic dysfunction on pulse rate aggregates in daily life. We are the first to study these relationships using real-life PPG data, rather than during controlled settings. We observed only a weak effect of autonomic dysfunction, as assessed through a patient-reported outcome (SCOPA-AUT), on maximum pulse rate. Interestingly, while the subjective measure showed a significant association, objective orthostatic hypotension measurements did not show a significant relationship with pulse rate parameters, despite numerically lower maximum pulse rates in the orthostatic hypotension group. Multiple factors could explain the weak effect overall. Maximum pulse rate is highly influenced by physical activity, which not only elevates the heart rate but also introduces motion artifacts that can lower PPG signal quality [32,33]. Although we adjusted for physical activity using the Physical Activity Scale for the Elderly (PASE), maximum pulse rate remains highly dependent on the amount and level of physical activity and the ability to capture high-quality PPG data during these activities. This is reflected in our findings, where the observed maximum pulse rates were relatively low compared to participants’ age-predicted maximum heart rates. Second, it is possible that the assessments for autonomic dysfunction in this study do not reliably capture all the physiological aspects of cardiac autonomic function. A more robust and objective alternative would be to use quantitative measures such as cardiac innervation imaging (123I-mIBG scintigraphy [34]). Given these limitations, future research should consider not only global pulse rate parameters but also zoom in on heart rate responses in more specific behavioral contexts, such as sleep or daily activities (e.g. during walking or other intense physical activities). Nocturnal analyses offer a promising avenue for research because reduced levels of movement during sleep will likely improve the PPG data quality. This would also allow for pulse rate variability measurements, which could yield more accurate assessments of cardiac autonomic function[35–37]. The merits of this approach remain to be formally demonstrated, as persons with PD can manifest a wide range of nocturnal movements, including dream enactment behavior and periodic leg movements during sleep [38].

### Strengths

A key strength of this study was the use of a large dataset collected from a representative population of people with early-stage PD through continuous real-world monitoring. Participants wore the smartwatch on average 22 hours per day, indicating strong adherence and consistent device use in daily life. This extensive dataset provides a comprehensive insight into pulse rate patterns in daily life of people with PD, including both daytime and nighttime recordings. Moreover, this study establishes a first link between free-living pulse rate data and autonomic dysfunction, offering new insights into autonomic regulation in PD. Another notable strength was the approach to data annotation and validation of pulse rate estimations. The careful annotations ensured the accuracy and reliability of the PPG signal quality, while the validation of the pulse rate estimation (using the smoothed pseudo Wigner-Ville distribution with alternative methods using an external dataset) reinforces the robustness of the methodology. Importantly, signal quality assessment explicitly accounted for periodic motion artifacts, such as tremors, by combining PPG and accelerometer signals. This multimodal approach allowed us to filter out segments with periodic disturbances, thereby improving the reliability of the pulse rate parameters. This supports the potential of PPG as a tool for monitoring pulse rate and autonomic function in people with PD. The Personalized Parkinson Project offers a unique opportunity to further study this concept, as we have digital data available for all participants for minimally up to 2 years (and up to 3 years for a sizeable subgroup) [11,39].

### Limitations

This study was not without limitations. The first limitation was the reliance on a self-administered questionnaire (SCOPA-AUT) to assess the severity of autonomic dysfunction. While this questionnaire is considered a gold-standard assessment for one’s autonomic symptoms, it remains inherently subjective and prone to high within-subject variability [40]. This could explain why we found no effect of autonomic dysfunction on resting pulse rate and only a weak effect on maximum pulse rate. Future studies should consider incorporating objective, quantitative measures of autonomic function, such as and cardiac innervation imaging using 123I-mIBG scintigraphy. In addition, the estimation of the effect of autonomic dysfunction on pulse rate parameters from observational data should be interpreted with caution, given its underlying assumptions. Potential residual confounding and imperfect measurement of key variables may have influenced the results. To increase transparency about these assumptions, we included a DAG, which may be further refined in future research. Another limitation is the lack of validation for the impact of filtering using ECG data. We did not explicitly assess whether segments with a high correlation between the spectral content of the accelerometer and PPG indeed resulted in erroneous pulse rate estimates when compared to ECG. Such an analysis would have helped to confirm that the high pulse rate estimates removed by filtering indeed reflected periodic motion artifacts, rather than physiological changes. However, our feature for periodic motion artifact detection was designed to identify segments with high accelerometer-PPG correlation. Therefore, we expect that filtering mainly removes implausible pulse rate estimates, while discarding only a minimal amount of real pulse rate data. Future research should confirm this by directly comparing filtered PPG data with ECG recordings. Finally, the potential impact of tremor on pulse rate estimation was only assessed at the group level using clinically based tremor assessments. Future work could benefit from integrating tremor and gait detection algorithms, particularly given the presence of low-frequency artifacts suggestive of possible gait-related motion. Such approaches would allow for per-segment analysis of periodic motion artifacts, providing a more precise evaluation of their effects on pulse rate measurements.

## Conclusion

This study presents a novel method for estimating pulse rate in the PD population. While the proportion of high-quality PPG data was high during nighttime, daytime data were more susceptible to motion contamination. Filtering periodic motion artifacts reduced overestimation of maximum pulse rate. An association was shown between lower daytime maximum pulse rate and more severe self-reported autonomic symptoms. Resting pulse rate showed no significant association with autonomic dysfunction, and no differences were observed with respect to objective orthostatic hypotension. These collective findings support the feasibility of using wearable PPG to assess cardiac autonomic function in PD and highlight the need for robust artifact-handling methods to advance digital biomarker development for this population.

## Acknowledgements

This work was financially supported by the Michael J Fox Foundation (grant #020425), the Dutch Research Council (grant #ASDI.2020.060 & grant #2023.010), the Dutch Research Council Long-Term Program (project #KICH3.LTP.20.006, financed by the Dutch Research Council, Verily, and the Dutch Ministry of Economic Affairs and Climate Policy), and by the Dutch Ministry of Economic Affairs (Topconsortium voor Kennis en Innovatie, Life Sciences & Health, grant #LSHM20090-H048). The Radboudumc Center of Expertise for Parkinson’s and Movement Disorders was supported by a center of excellence grant from the Parkinson’s Foundation.

## Author contributions

Research project (1): A. Conception, B. Organization, C. Execution;

Statistical Analysis (2): A. Design, B. Execution, C. Review and Critique;

Manuscript Preparation (3): A. Writing of the first draft, B. Review and Critique;

K.I.V.: 1A, 1B, 1C, 2A, 2B, 3A.

L.J.W.E.: 1A, 1B, 2A, 2C, 3B.

T.L.: 1A, 2A, 2C, 3B.

J.P.Y.: 1A, 2C, 3B.

M.A.L.: 1A, 2C, 3B.

K.C.H.: 2C, 3B.

S.S.: 2C, 3B.

L.S.L: 2C, 3B

B.R.B.: 1A, 1B, 2C, 3B.

M.A.B.: 1A, 2C, 3B.

J.T.: 1A, 1C, 2A, 2C, 3B.

