## Supplementary Materials for "Heart rate monitoring using wrist photoplethysmography in Parkinson disease: feasibility and relation with autonomic dysfunction"

### 1 Supplementary methods

#### 1.1 Development of PPG morphology classifier

The goal of the classifier was to distinguish high quality PPG data from poor quality, based on short (6s) PPG segments derived from manually annotated 1-minute epochs. The classifier was trained on handcrafted features extracted from these segments and evaluated using a nested cross-validation procedure to ensure generalizability.

##### 1.1.1 Annotation pipeline

The annotation pipeline relied on visual inspection by experts, supported by consistent signal visualization. First, PPG recordings were divided into 1-minute epochs. Then, filtered signals were visualized alongside their spectrograms using a custom graphical user interface (GUI). Annotations were performed independently by two trained investigators.

###### 1.1.1.1 Annotation data set

A representative sample of 20 PPP subjects was selected from the data set to create the annotation set. Stratification was performed based on the following characteristics: sex, age and resting tremor scores (UPDRS III: Motor examination in OFF state). From each subject, 1-minute PPG epochs were selected from two timepoints: week 1 and week 52. For each week, annotation was performed on the first full 24h of the PPG recording (1440 epochs). This approach ensured that the total annotation set (57600 epochs) included variability in both temporal aspects and individual signal characteristics.

###### 1.1.1.2 Data quality labeling

All PPG recordings were divided into non-overlapping 1-minute epochs, which allowed for sufficient context for reliable visual interpretation of PPG morphology while remaining also practical for large-scale annotations. We determined high quality PPG data as the typical waveform as described in literature^1–3^. Each epoch was assigned to one of the initial four quality categories, defined by the proportion of the signal displaying the typical morphology:

- Label 1 – Very high quality: ≥95% of the epoch
- Label 2 – Moderate quality: 50-95%
- Label 3 – Low quality: 10-50%
- Label 4 – Very poor quality: <10%

###### 1.1.1.3 Annotation protocol

Annotations were performed using a custom GUI in MATLAB. Each 1-minute epoch was shown with:

- Time-domain PPG signal (filtered, fixed amplitude scale)
- Spectrogram (computed using pspectrum, 3s time resolution, 50% overlap)
- Timestamp (absolute time in the recording)

Spectrograms were min-max normalized to a range between 0 and 1 for better comparability of the dominant frequencies. Investigators underwent a training phase using three independent 24h datasets and proceeded to annotate after reaching inter-rater agreement (Cohen’s kappa > 0.8).

##### 1.1.2 Feature extraction

For the development of the classifier in this research, we only used the epochs annotated as label 1 (very high quality) or label 4 (very poor quality). These epochs were subdivided into 6s non-overlapping segments. From each 6s PPG segment, a set of 10 time- and frequency-domain features was constructed. The following 10 features were calculated:

- Standard deviation
- Mean amplitude of the absolute PPG signal
- Median amplitude of the absolute PPG signal
- Skewness
- Kurtosis
- Dominant frequency: calculated using Welch’s method with a 3s Hann window and 50% overlap.
- Relative power: power within ± 0.2 Hz around the dominant frequency, normalized to the total power
- Spectral entropy: computed as the Shannon entropy of the normalized power spectral density
- Signal-to-noise ratio: ratio of the variance of the absolute signal (signal variance) to the variance of the signal (noise variance), as described by Elgendi et al.^2^
- Autocorrelation peak: highest non-zero-lag peak of the autocorrelation function

##### 1.1.3 Training of the classifier

###### 1.1.3.1 Classifier

A Logistic Regression (LR) classifier was selected due to its interpretability and effectiveness for binary classification.

###### 1.1.3.2 Hyperparameter optimization

For hyperparameter optimization, a grid search approach was used to identify the best configuration of hyperparameters for the LR classifier. The following hyperparameters were tuned:

- Regularization strength (λ): varied over a logarithmic range, including 0 and log-spaced values from 10^-6^ and 10^0^. The regularization method was fixed to lasso (L1 regularization)
- Solver: the optimization process explored two solvers:
  - Stochastic Gradient Descent (SGD)
  - Sparse Reconstruction by Separable Approximation (SpaRSA)

###### 1.1.3.3 Leave-One-Out Cross-Validation (LOOCV)

In the LOOCV, the dataset split such that one subject is held out for testing, and the remaining subjects are used for training. In the inner loop, hyperparameters are optimized through grid search based on the highest average performance across the training set. In the outer loop, the model is evaluated on the held-out subject using the optimal hyperparameters. The performance is averaged across all iterations. Finally, hyperparameters are selected based on the average performance of the held-out validation data, and the model is retrained on the full dataset using the optimal configuration.

##### 1.1.4 Results

The LR classifier achieved high performance in distinguish high-quality from poor quality segments. Supplementary Table 2 shows the classification metrics averaged over all LOOCV iterations.

**Supplementary Table 1: Performance metrics of the logistic regression classifier evaluated using nested leave-one-out cross-validation (LOOCV) on 6s PPG segments. Metrics are averaged across all LOOCV iterations.**

| Metric | Mean (± SD) |
| --- | --- |
| Accuracy | 98.4 (1.0) |
| Sensitivity | 98.2 (1.4) |
| Specificity | 98.5 (1.3) |
| F1-score | 98.1 (1.1) |
| Precision | 97.9 (1.7) |

The best performance of the final LR classifier was achieved using a LR classifier with a regularization parameters of 0.0001 using Lasso regularization and the SpaRSA solver.

**Supplementary Table 2: Final hyperparameter configuration for the logistic regression classifier after nested leave-one-out cross-validation.**

| Hyperparameter | Mean (± SD) |
| --- | --- |
| Regularization strength (λ) | 0.0001 |
| Regularization type | Lasso (L1) |
| Solver | SpaRSA |

#### 1.2 Periodic motion artifact removal

##### 1.2.1 Calculation of Relative Power

To identify periodic artifacts in the PPG signal caused by periodic motion artifacts, we employed a method to calculate the window-based relative power in the accelerometer signal. This process involved the following steps (also illustrated in Supplementary Figure 1):

1. **Frequency Analysis**: We performed the Welch’s method on both the PPG and accelerometer signals to obtain the frequency distribution. We defined the dominant frequency as the frequency at which the signal has the highest power.
2. **Frequency Band Selection**: For each window, we focused on the dominant PPG frequency (±0.05 Hz) and its first harmonic (±0.05 Hz). The first harmonic is determined as twice the dominant frequency and is included to capture higher-order periodic components.
3. **Power Calculation**: Using the PSD estimates from the Welch’s method, we calculated the power within these frequency bands for the accelerometer signal. The power within a frequency band is the integral of the PSD over that band, which we computed using the trapezoidal rule.
4. **Relative Power**: The relative power is defined as the ratio of the power within the specified frequency band to the total power in the accelerometer signal. This ratio quantifies the proportion of the signal's energy that is concentrated at the dominant PPG frequency and its harmonic.

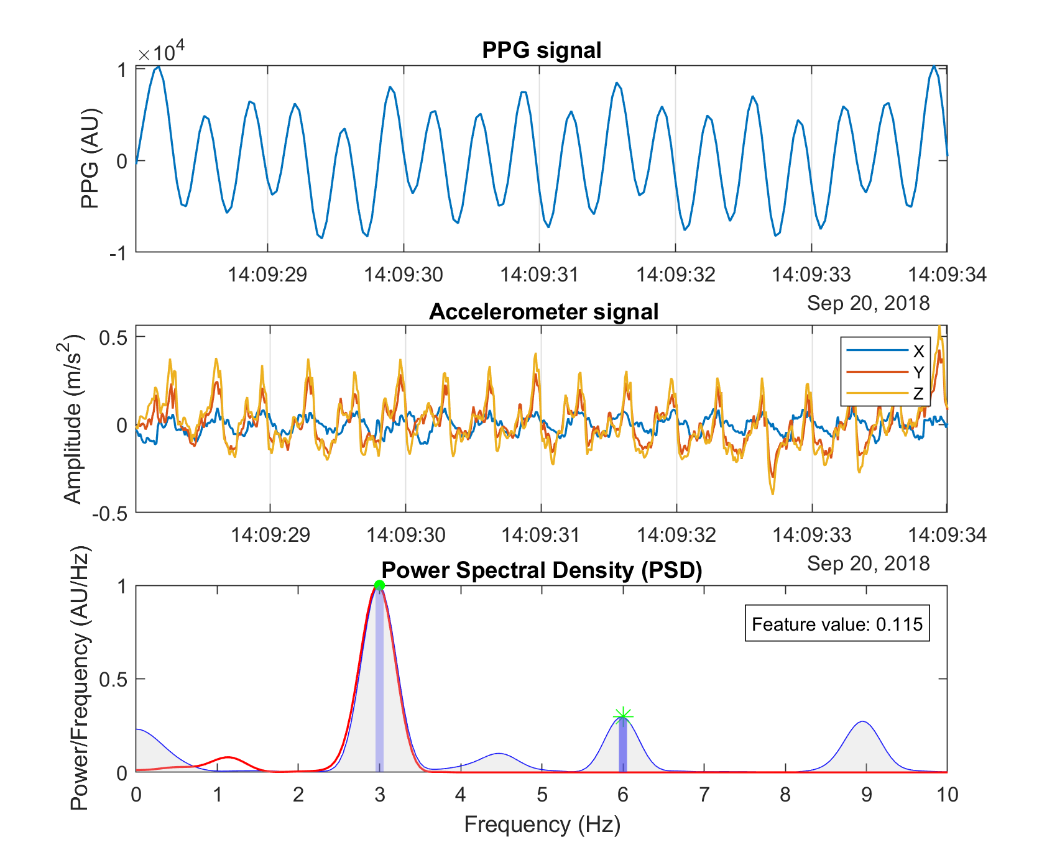

**Supplementary Figure 1: Schematic illustration of the relative power feature of the accelerometer. Upper panel: PPG signal, middle panel: three axes of the accelerometer, lower panel: visualization of the relative power feature. Frequency content of both signals is obtained using Welch’s method. Thereafter, the dominant frequency (green dot) and first harmonic (green star) of the PPG signal are detected. Around both frequencies (±0.05 Hz) we calculate band powers (blue areas) using trapezoidal rule and calculate the relative power compared to the total power (grey area). In this example the feature value is 0.115.**

##### 1.2.2 Threshold Selection

The threshold for classifying a window as an artifact was determined through empirical analysis of the annotation data set. The steps involved were:

1. **Training Dataset Analysis**: We analyzed the same data set as was used for developing the LR classifier to assess PPG morphology. We used every 6s segment which was classified as high-quality PPG based on this classifier for all 20 subjects in the first study weeks. This resulted in 5.2 million segments.
2. **Relative Power Distribution**: We calculated the relative power for all segments in the training dataset and examined the total distribution of relative power values (Supplementary Figure 2).
3. **Threshold Determination**: We set a threshold of 0.10 for the relative power. This threshold was chosen based on total distribution as shown in Supplementary Figure 2. While the overall distribution resembles an exponential decay, we observed a deviation from this trend around 0.10, where the feature values occur more frequently than expected. This suggests that values above this threshold may represent a qualitatively different subset of data, making 0.10 a reasonable and data-driven cut-off.

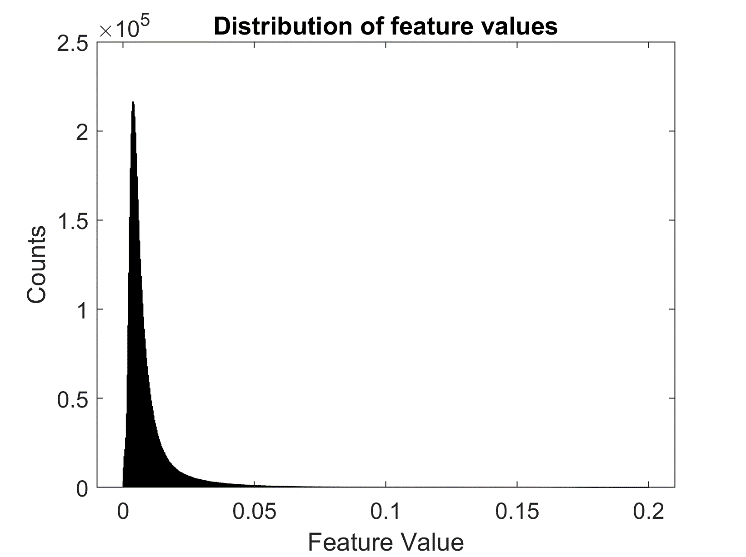

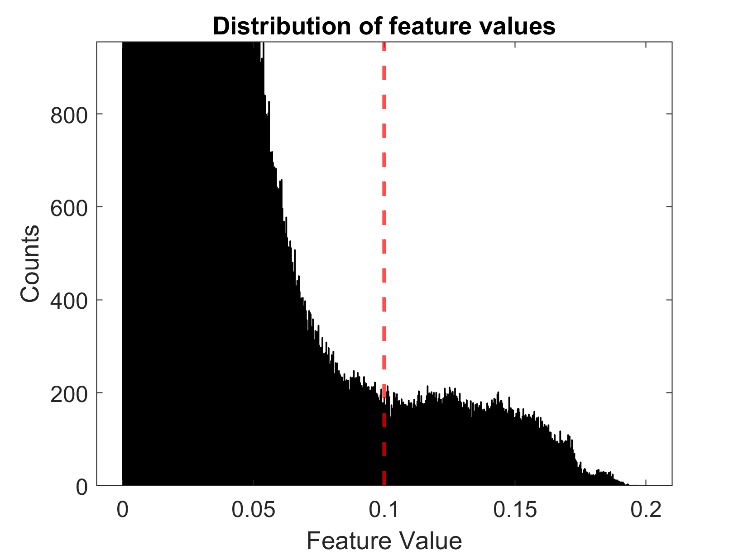

**Supplementary Figure 2: The left panel shows the overall distribution of the relative power values, following an exponential decay. The right panel provided a zoomed-in view showing a notable deviation from the exponential decay trend around the feature value of 0.10. Based on this deviation, a threshold of 0.10 was established for classifying artifacts.**

#### 1.3 Signal quality classification

The LR classifier produced posterior probabilities, where higher probabilities indicated a higher likelihood of the PPG signal being of high quality, characterized by clear pulse waves with discernible peaks. For each 1-second window, the mean posterior probability was calculated, and a window was classified as high-quality if its mean probability exceeded 0.5. Accelerometer-derived binary labels were used to refine the classification by detecting periodic motion artifacts. A label of 0 indicated substantial motion artifact (above the threshold). If the majority of accelerometer labels in a 1-second window were 0, the overall signal quality label was set to 0, overriding the PPG morphology classification. For each 1-sec window, the mean posterior probability was calculated. A window was classified as high-quality if its mean posterior probability exceeded 0.5. The accelerometer-derived labels binary were used to refine the signal quality classification by detecting periodic motion artifacts. A label of 0 indicated substantial motion artifact (exceeding the threshold). If the majority-voted accelerometer label for a 1-sec window was equal to 0, the overall signal quality label was set to 0, overriding the PPG morphology classification.

#### 1.4 Validation of Pulse Rate Estimation

To evaluate the most accurate approach for heart rate estimation from photoplethysmography signals, we compared six different methods, including four time-frequency analysis techniques and two beat-detection algorithms. While beat-detection methods directly identify individual pulses, time-frequency methods offer a continuous representation, which can be more beneficial In cases of noisy signals. To ensure reliable comparisons, we applied the signal quality algorithm to extract high-quality PPG segments of at least 30s from an external dataset, containing synchronized PPG and ECG recordings during free-living activities. Pulse rate was then estimated on these segments for every 2s, using each of the six methods. By comparing both approaches, we aimed to assess their strengths and determine which method provides the most reliable estimates under ambulatory conditions.

##### 1.4.1 Validation dataset

The performance of each method was assessed using the PPG DaLiA dataset^4^_,_ which contains synchronized PPG and electrocardiogram (ECG) recordings from subjects engaged in daily activities. ECG-derived heart rate of every 2s (8s windows, 6s overlap) served as the reference standard.

##### 1.4.2 Time frequency methods

Time-frequency distributions (TFDs) provide a representation of the PPG signal’s spectral content over time, allowing for pulse rate estimation by identifying the dominant frequency component in the expected physiologically heart rate range (40-180 bpm). The following four TFD methods were considered and implemented in MATLAB:

**Short-Time Fourier Transform (STFT):** The STFT partitions the signal into windows and applies the Fourier transform to each segment^5^_._ This approach provides a straightforward way to analyze frequency variations. However, its resolution is limited by the fixed window length, leading to trade-offs between time and frequency precision. A shorter window improves the temporal resolution but reduces frequency precision while a longer window has the opposite effect^6^. To match the ECG labels, we used 8s windows with 6s overlap.

**Continuous Wavelet Transform (CWT):** The CWT decomposes the signal into wavelet coefficients across multiple scales, capturing both transient and oscillatory components^5^_._ This multi-scale approach provides adaptability to pulse rate variations, but its resolution depends on the choice of wavelet function and may be affected by noise in non-stationary signals. For this specific analysis, the Generalized Morse Wavelet was implemented using the in-build *cwt* function.

**Wigner-Ville Distribution (WVD):** The WVD provides a high-resolution time-frequency representation by computing the time-dependent spectral energy of the signal. This method offers precise frequency localization but is highly susceptible to cross-term interference when multiple frequency components are present.^5^

**Smoothed Pseudo Wigner-Ville Distribution (SPWVD):** The SPWVD is a modification of the WVD that incorporates time and frequency smoothing to mitigate the presence of cross-terms, which can obscure the true spectral content of the PPG signal.^5^ The smoothing functions improve robustness against noise, making it particularly effective in tracking pulse rate variations during noisier segments. Compared to standard WVD, SPWVD tries to maintain a high-frequency resolution without suffering from cross-terms. For the WVD and SPWVD, a fast and memory-efficient implementation of both algorithms was used.^7^ The SPWVD was smoothed using 1s hamming window in the time domain and a 8s hamming window in the frequency domain, determined empirically.

##### 1.4.3 Beat-detection algorithms

In addition to time-frequency distribution methods, we also evaluated two robust beat-detection algorithms^8^, which aim to identify individual heartbeats from PPG signals and derive 2s pulse rate estimates based on inter-beat-intervals. To do this, we detected peaks within 8s windows with a 6s overlap. The two algorithms were:

**Multi-Scale Peak and Trough Detection (MSPTD):** This method detects peaks and onsets at multiple scales in the PPG signal to enhance robustness against noise and artifacts.

**Adapted onset detector (qPPG):** This method detects heartbeats by analyzing the steepest upward slopes in the PPG signal. It uses a slope function over a short window and applies an adaptive threshold to identify heartbeats.

##### 1.4.4 Evaluation metrics

Accuracy of the different methods was assessed using the following metrics:

- Mean absolute error: The mean of the absolute differences between the 2s pulse rate estimates from PPG and the reference ECG label. This provides a general indication of accuracy.
- Median absolute error: The median of the absolute differences between the 2s pulse rate estimates from PPG and the reference ECG label. This provides an indication of accuracy without influence of outliers.
- Percentage of windows absolute error <2 BPMs: To evaluate the proportion of highly accurate estimates, reflecting reliability in estimating the correct heart rate.
- Percentage of windows absolute error > 5 BPM: To assess the frequency of large errors, indicating robustness against significant inaccuracies in heart rate estimations.

##### 1.4.5 Results

**Supplementary Table 3: Evaluation metrics of the validation for six different methods to perform heart rate estimation on PPG signals. CWT = Continuous Wavelet Transform, MSPTD = Multi-Scale Peak and Trough Detection, qPPG = Adapted Onset Detector, SPWVD = Smoothed-Pseudo Wigner-Ville Distribution, STFT = Short-Time Fourier Transform, WVD = Wigner-Ville Distribution.**

|  | Time frequency analyses | | | | Beat detectors | | |
| --- | --- | --- | --- | --- | --- | --- | --- |
| Method | **STFT** | **CWT** | **WVD** | **SPWVD** | | **qPPG** | **MSPTD** |
| Mean absolute error (SD) | 1.34  (2.30) | 1.12  (2.03) | 1.34  (2.48) | 0.87  (2.11) | | 1.98  (4.31) | 1.38  (2.22) |
| Median absolute error (IQR) | 0.80  (0.37 – 1.60) | 0.75  (0.35 – 1.33) | 0.67  (0.30 – 1.36) | 0.44  (0.20 – 0.89) | | 0.93  (0.41 – 1.86) | 0.90  (0.40 – 1.71) |
| % absolute error < 2 | 82.0 | 88.4 | 84.3 | 92.5 | | 77.1 | 79.9 |
| % absolute error > 5 | 3.0 | 1.1 | 4.8 | 1.1 | | 5.5 | 2.4 |

##### 1.4.6 Conclusion

The results suggest that time-frequency approaches are more reliable than the evaluated beat detection algorithms for heart rate estimation from PPG. Among these methods, the SPWVD provided the most accurate pulse rate estimates, with the lowest overall error metrics and the highest proportion of accurate estimates. This highlights SPWVD as the most robust choice for analysis.

##### 1.4.7 Example of SPWVD

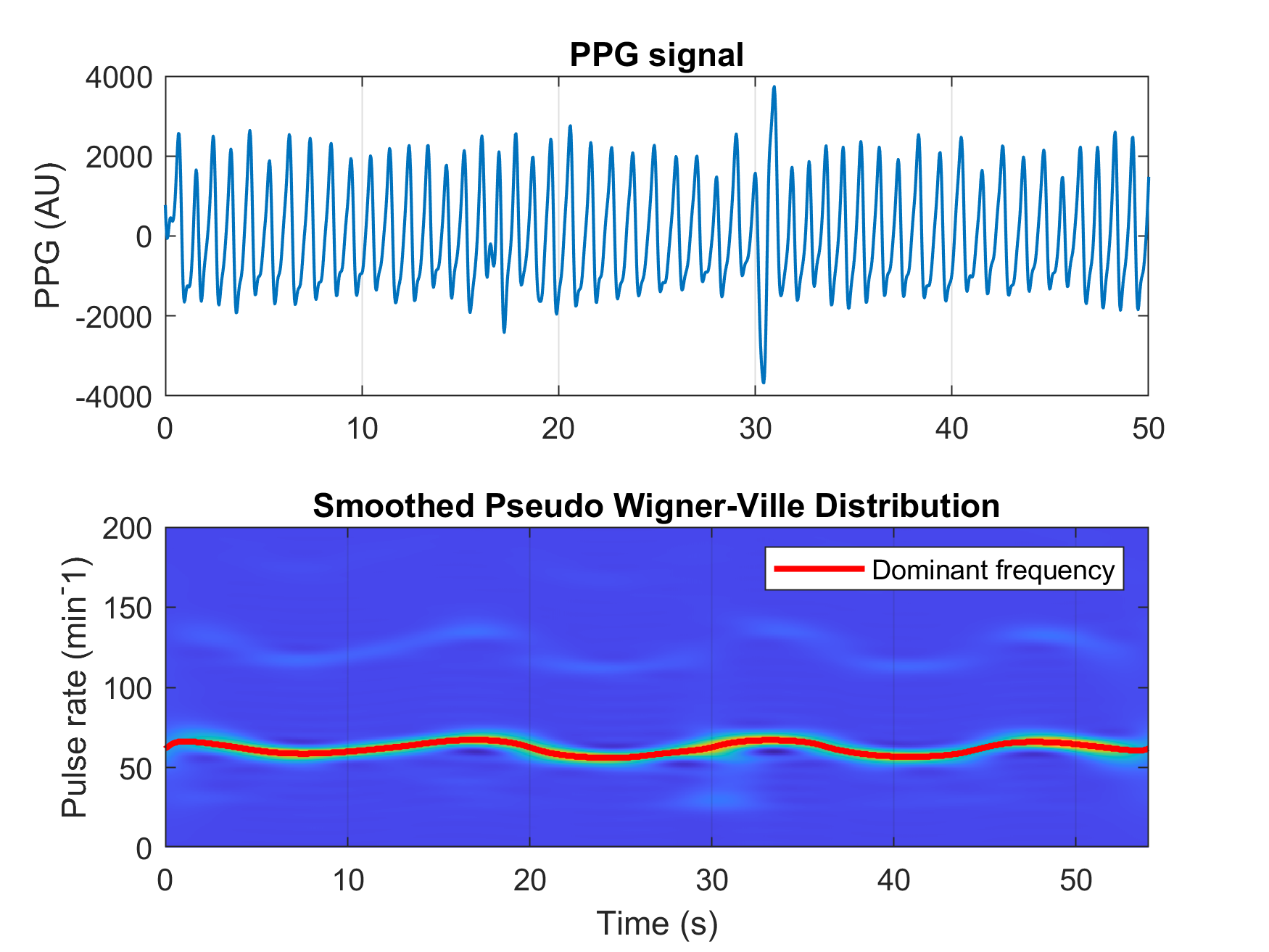

**Supplementary Figure 3: Example of the smoothed pseudo Wigner-Ville distribution (SWPVD) applied to a the PPG signal segment. The upper panel shows the preprocessed PPG segment, while the lower panel displays its SPWVD. The red line indicates the dominant frequency at each time point. Pulse rate was estimated for every two-second window by averaging the dominant frequencies within that window.**

#### 1.5 Directed acyclic graph pulse rate analyses

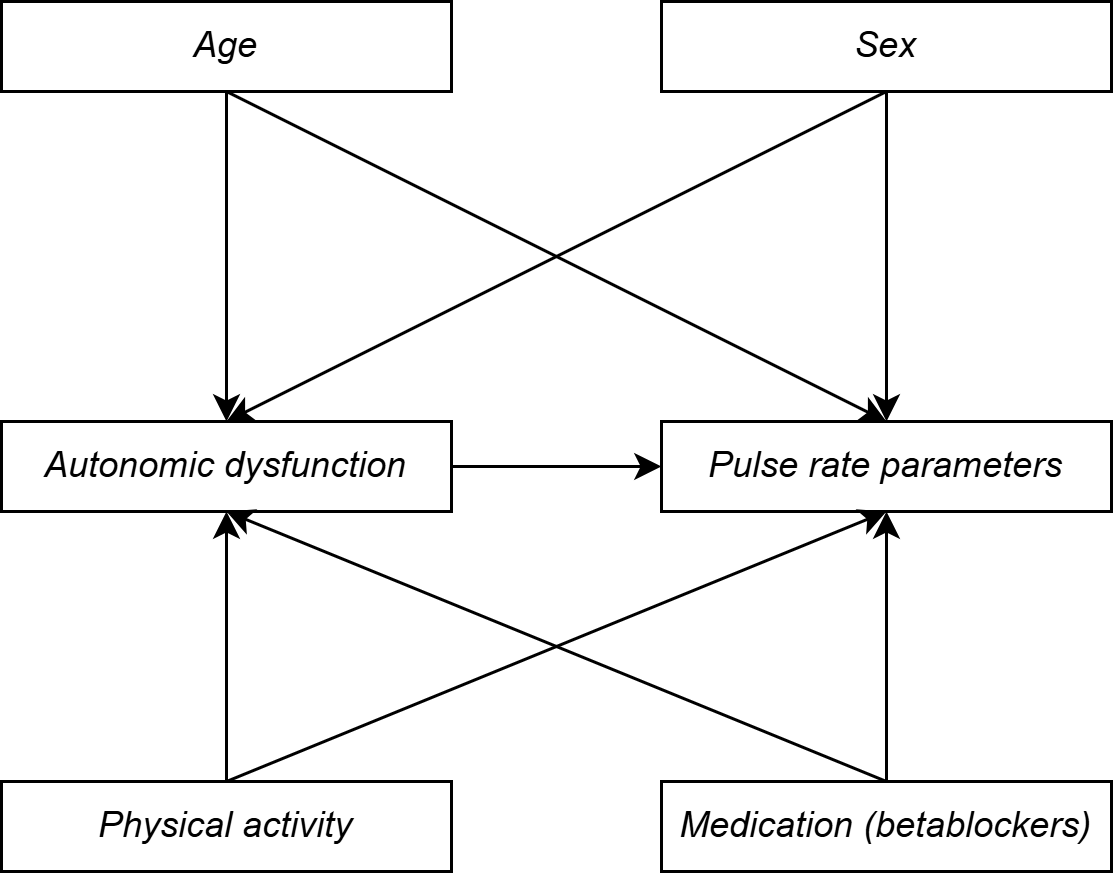

**Supplementary Figure 4: Directed Acyclic Graph illustrating the known relationships between covariates and pulse rate parameters and autonomic dysfunction. Arrows indicate the direction of influence. Age, sex, physical activity and medication (specifically beta-blockers) are included as covariates in this study model to adjust for their potential confounding effects.**

### 2 Supplementary Results

#### 2.1 Data quality week 1

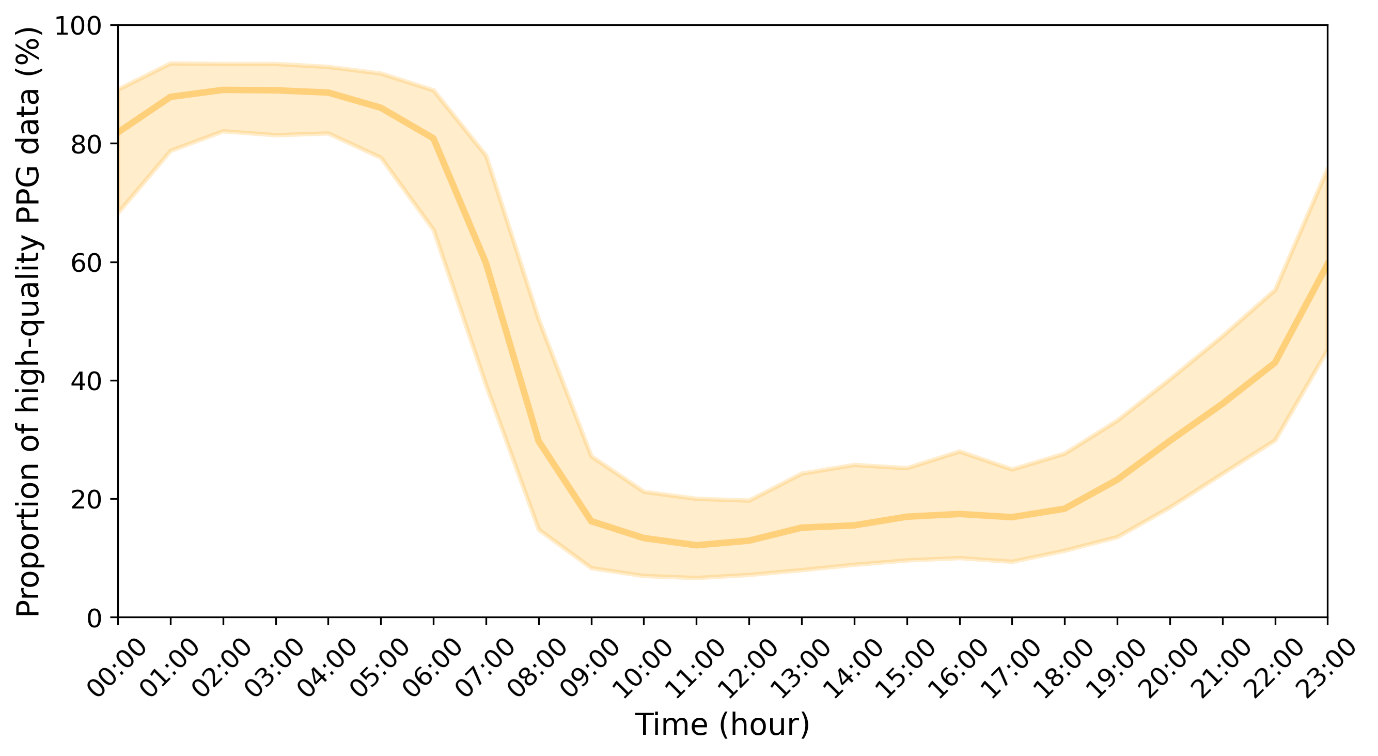

**Supplementary Figure 5: Proportion of high-quality PPG data across the circadian cycle in the second study week. Data are represented as medians + IQR.**

#### 2.2 Influence of periodic artifact removal (step 2)

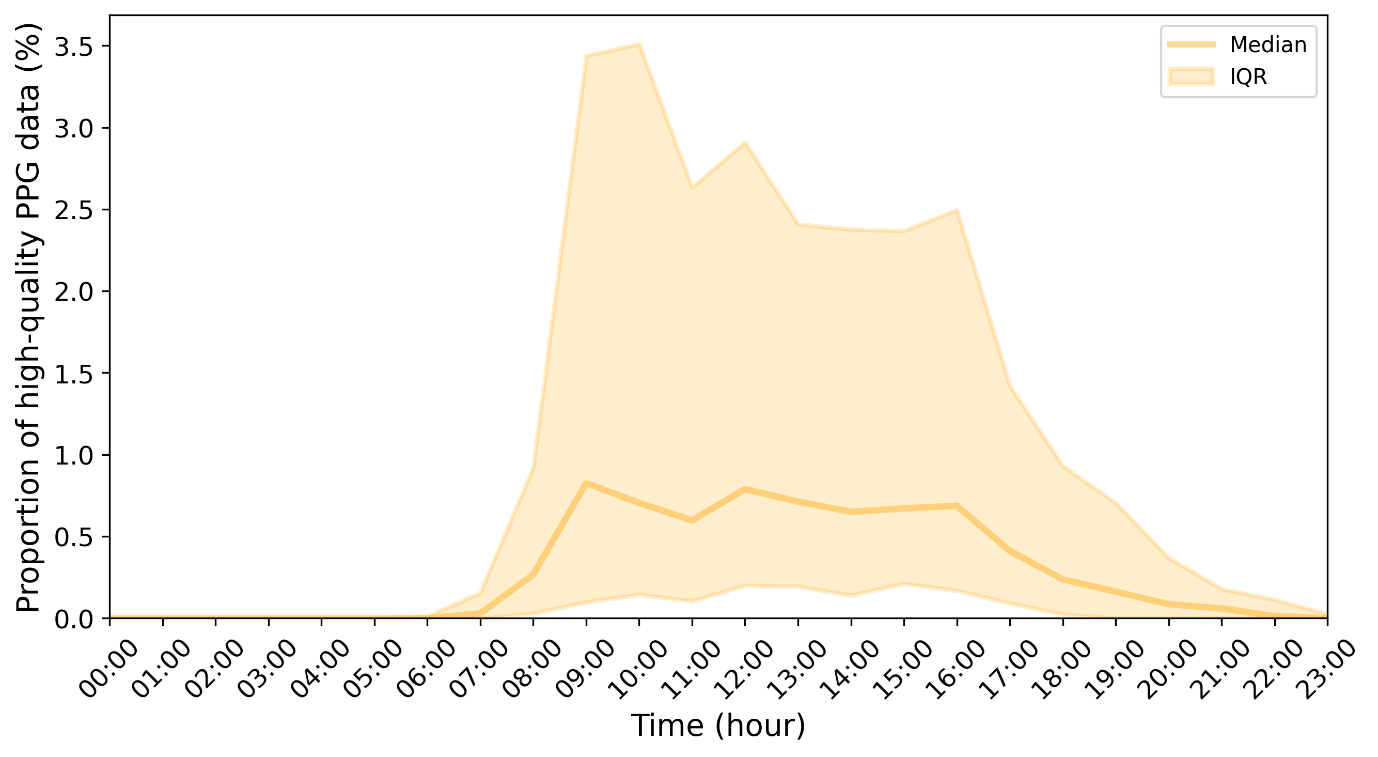
**Supplementary Figure 6: Removal of the proportion of high-quality PPG data after applying periodic artifact removal (step 2) compared to only step 1 across the circadian cycle in the first study week. Data are represented as medians + IQR.**

#### 2.3 Impact of dyskinesia on signal quality

|  | **No tremor (n=293)** | **Mild tremor**  **(n=101)** | **Severe tremor (n=50)** | **All**  **(n=444)** |
| --- | --- | --- | --- | --- |
| ***Week 0*** |  |  |  |  |
| Data quality day (%) | 28.95  [24.33, 36.29] | 29.68  [23.98, 35.67] | 30.72  [22.36, 35.86] | 29.17  [24.01, 35.90] |
| Data quality night (%) | 85.90  [79.24, 90.57] | 86.73  [78.68, 90.46] | 85.38  [79.73, 90.74] | 86.08  [79.30, 90.57] |
| Step 1 removal: Atypical morphology (PPG) (%) | 70.78  [63.48, 75.39] | 70.28  [64.23, 75.99] | 69.18  [64.07, 77.59] | 70.58  [63.97, 75.88] |
| Step 2 removal: Periodic movement (accelerometer) (%) | 0.11  [0.04, 0.24] | 0.08  [0.02, 0.23] | 0.08  [0.03, 0.20] | 0.10  [0.03, 0.22] |
| ***Week 1*** |  |  |  |  |
| Data quality day (%) | 29.48  [24.38, 36.50] | 28.97  [24.66, 34.86] | 27.34  [19.94, 37.12] | 29.01  [23.82, 36.19] |
| Data quality night (%) | 86.37  [79.39, 90.70] | 87.66  [79.19, 91.47] | 85.16  [80.65, 91.40] | 86.34  [79.37, 91.03] |
| Step 1 removal: Atypical morphology (PPG) (%) | 70.48  [63.20, 75.41] | 71.01  [65.12, 75.24] | 72.24  [62.19, 79.84] | 70.76  [63.46, 75.89] |
| Step 2 removal: Periodic movement (accelerometer) (%) | 0.12  [0.04, 0.29] | 0.08  [0.02, 0.22] | 0.09  [0.03, 0.26] | 0.11  [0.03, 0.28] |

***Supplementary Table 3: Data quality across different tertiles of different rest tremor severities. Data are presented as median + IQR.***

***Supplementary Table 4: Data quality across tertiles with different dyskinesia severities. Data are presented as median + IQR.***

|  | **No dyskinesia (n = 367)** | **Mild dyskinesia**  **(n = 35)** | **Severe dyskinesia**  **(n = 41)** | **All**  **(n=444)** |
| --- | --- | --- | --- | --- |
| ***Week 0*** |  |  |  |  |
| Data quality day (%) | 29.27  [24.03, 35.91] | 30.18  [25.64, 37.20] | 28.36  [21.39, 34.83] | 29.17  [24.01, 35.90] |
| Data quality night (%) | 86.16  [79.15, 90.54] | 84.56  [82.11, 89.61] | 85.92  [78.52, 91.07] | 86.08  [79.30, 90.57] |
| Step 1 removal: Atypical morphology (PPG) (%) | 70.42  [64.01, 75.81] | 69.79  [62.48, 74.29] | 71.43  [64.50, 78.45] | 70.58  [63.97, 75.88] |
| Step 2 removal: Periodic movement (accelerometer) (%) | 0.10  [0.03, 0.23] | 0.11  [0.03, 0.24] | 0.10  [0.03, 0.20] | 0.10  [0.03, 0.22] |
| ***Week 1*** |  |  |  |  |
| Data quality day (%) | 29.16  [24.38, 36.39] | 28.69  [24.51, 35.58] | 28.52  [22.32, 34.45] | 29.01  [23.82, 36.19] |
| Data quality night (%) | 86.41  [79.24, 91.17] | 86.05  [80.09, 89.78] | 87.39  [82.73, 90.98] | 86.34  [79.37, 91.03] |
| Step 1 removal: Atypical morphology (PPG) (%) | 70.51  [63.31, 75.46] | 71.29  [64.29, 75.41] | 70.98  [65.42, 77.58] | 70.76  [63.46, 75.89] |
| Step 2 removal: Periodic movement (accelerometer) (%) | 0.12  [0.03, 0.29] | 0.08  [0.04, 0.13] | 0.10  [0.03, 0.18] | 0.11  [0.03, 0.28] |

#### 2.4 Influence of periodic artifact removal (step 2) on pulse rate estimates

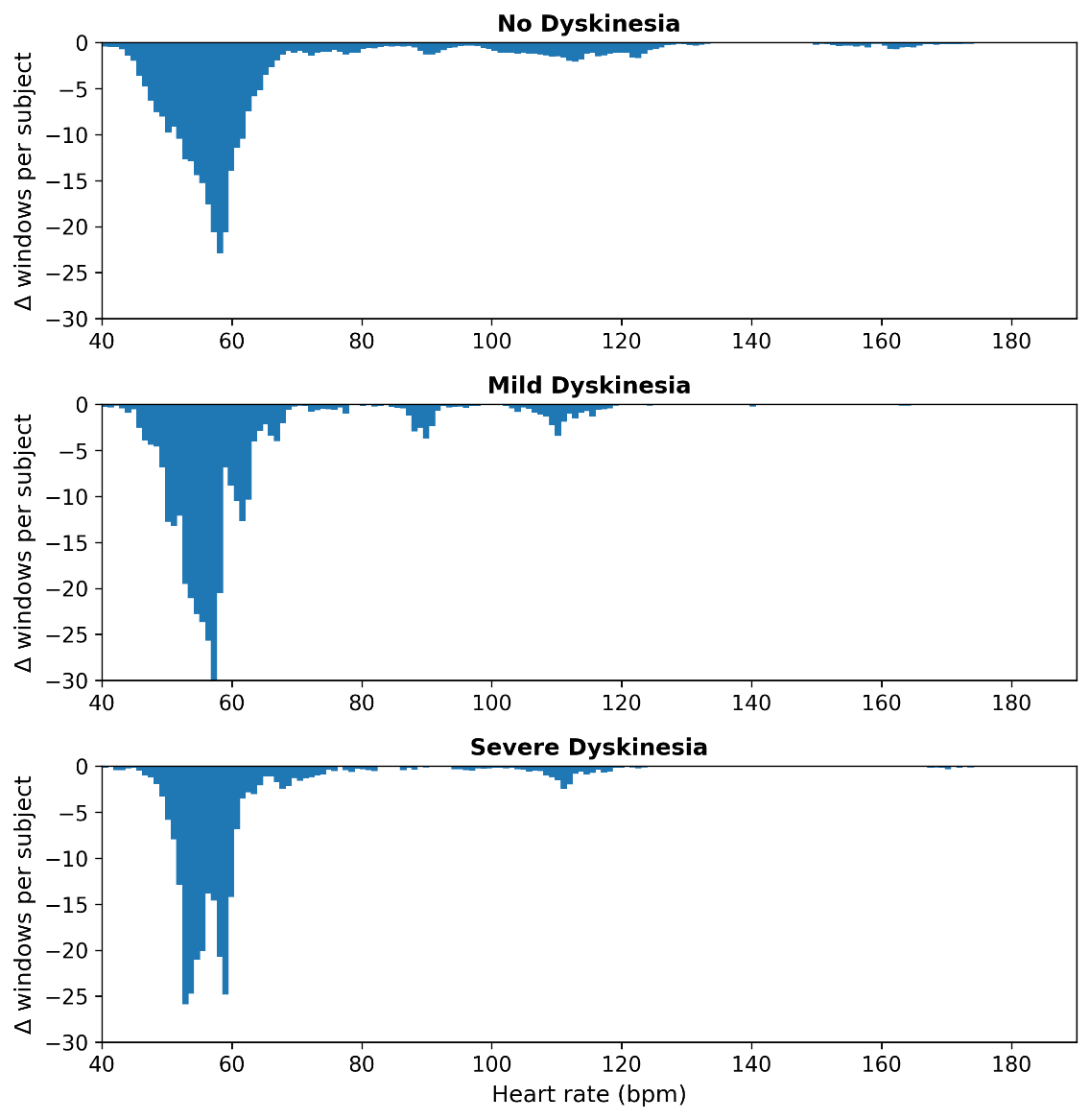

**Supplementary Figure 7: Removed pulse rate estimates by applying step 2 of the signal quality algorithm, for the different dyskinesia tertiles. The data show the mean change in counts per subject for each pulse rate value. There is no association between removed pulse rate estimates and dyskinesia severity.**

#### 2.5 Pulse rate parameters - descriptives

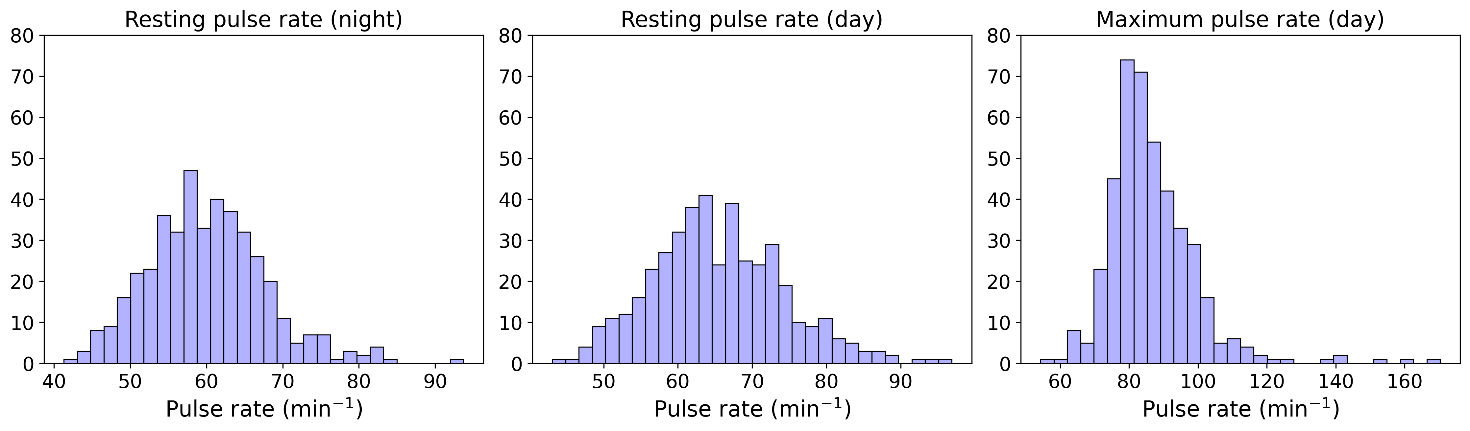

**Supplementary Figure 8: Distributions of the three different pulse parameters in the first study week across the study population.**

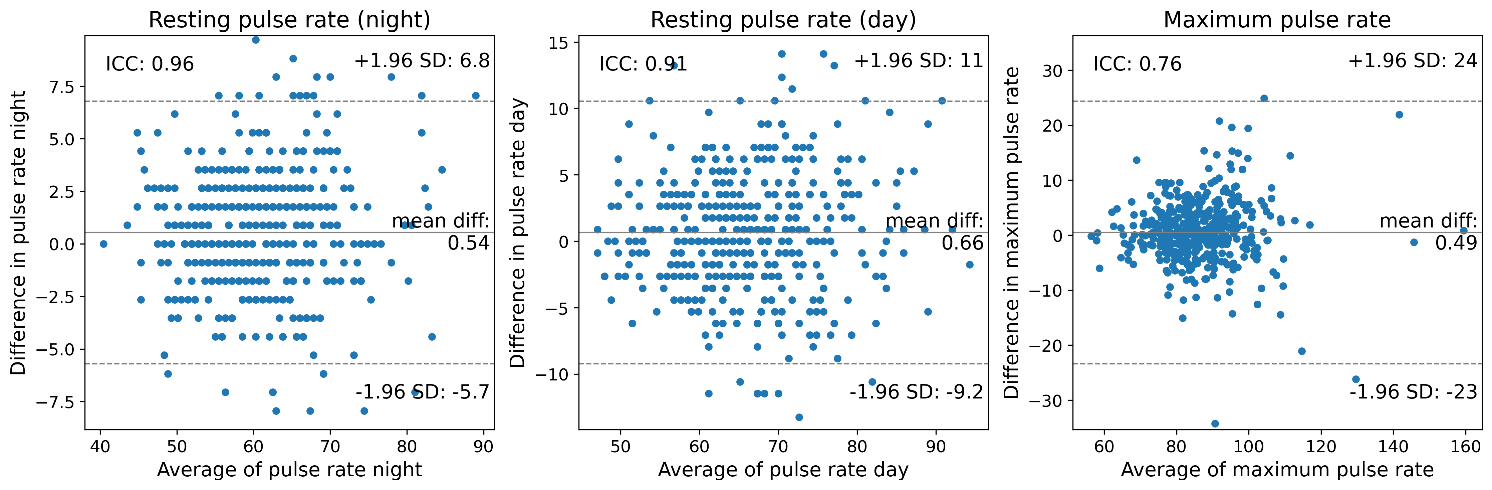

**Supplementary Figure 9: Bland-Altman plots of the pulse rate parameters in the first two study weeks with the intraclass correlation coefficient.**

#### 2.6 Influence of periodic artifact removal (step 2) on pulse rate parameters

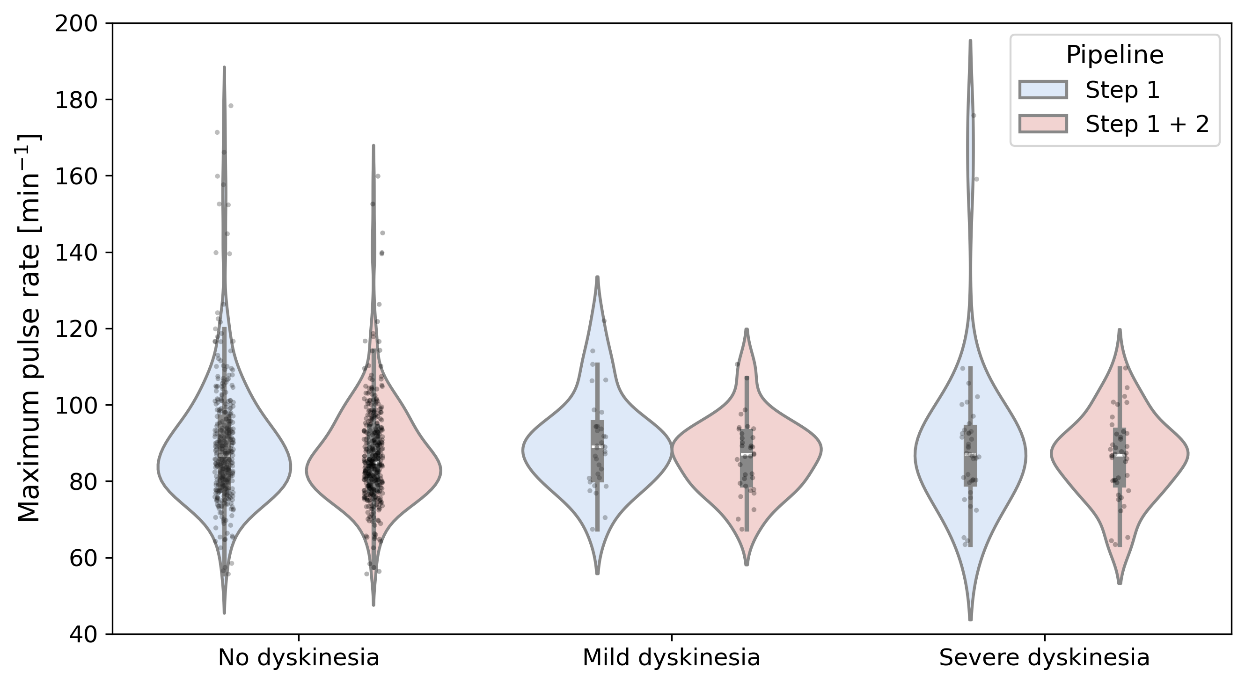

**Supplementary Figure 10: Violins of the maximum pulse rate in relation to dyskinesia severity (MDS-UPDRS 4.1 + 4.2) in the first study week. The different colors represent the maximum pulse rate when assessing solely PPG morphology (blue) or also incorporating periodic artifact removal using the accelerometer (red). Filtering for periodic motion artifacts does not significantly affect the aggregated maximum pulse rate at the group level, but does influence estimates at the individual level in any dyskinesia group.**

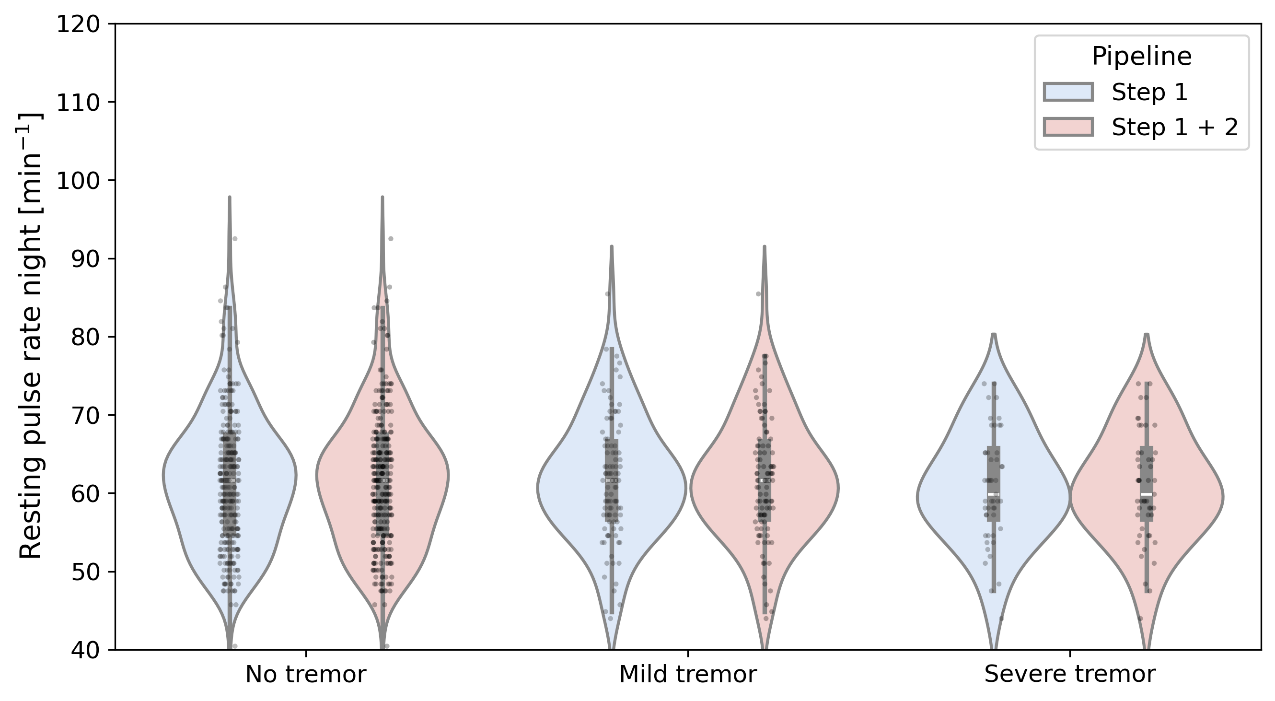

**Supplementary Figure 11: Violins of the resting pulse rate during the night in relation to tremor severity (MDS-UPDRS 3.17) in the first study week. The different colors represent the resting pulse rate when assessing solely PPG morphology (blue) or also incorporating periodic artifact removal using the accelerometer (red). Filtering for periodic motion artifacts does not affect the aggregated resting pulse rate in any tremor group.**

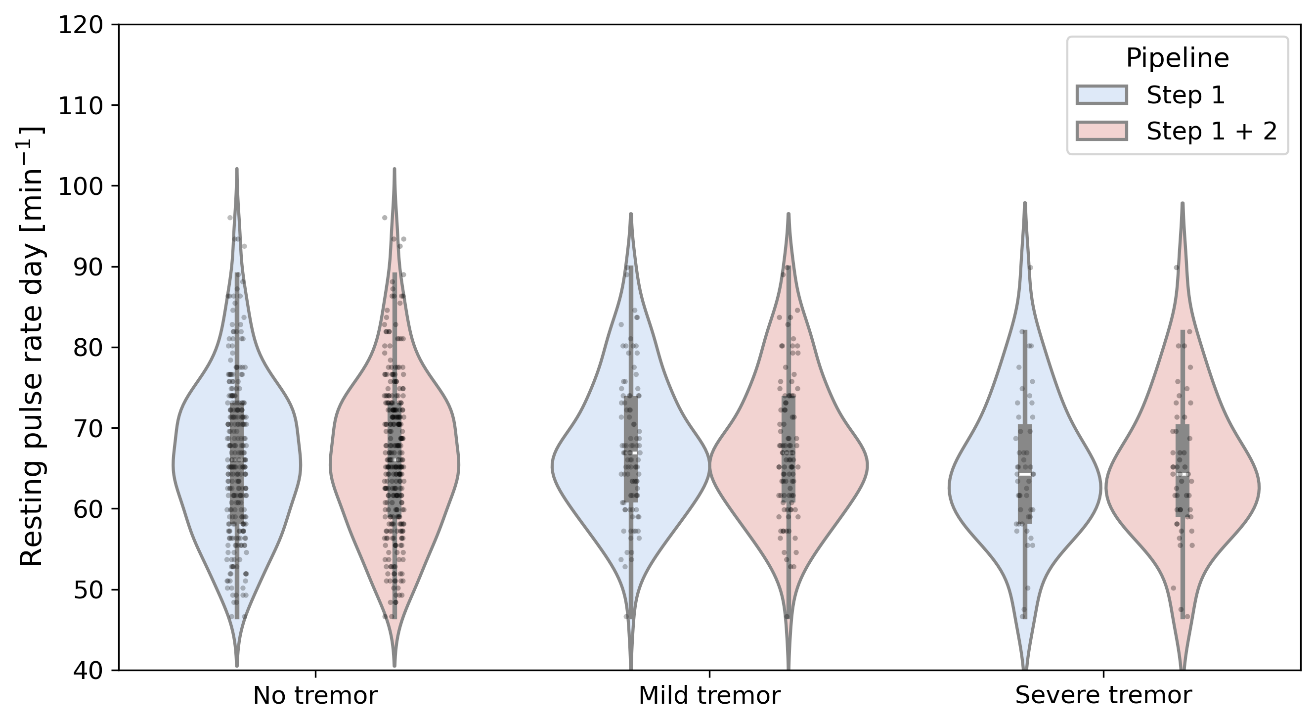
**Supplementary Figure 12: Violins of the resting pulse rate during the day in relation to tremor severity (MDS-UPDRS 3.17) in the first study week. The different colors represent the resting pulse rate when assessing solely PPG morphology (blue) or also incorporating periodic artifact removal using the accelerometer (red). Filtering for periodic motion artifacts does not affect the aggregated resting pulse rate in any tremor group.**

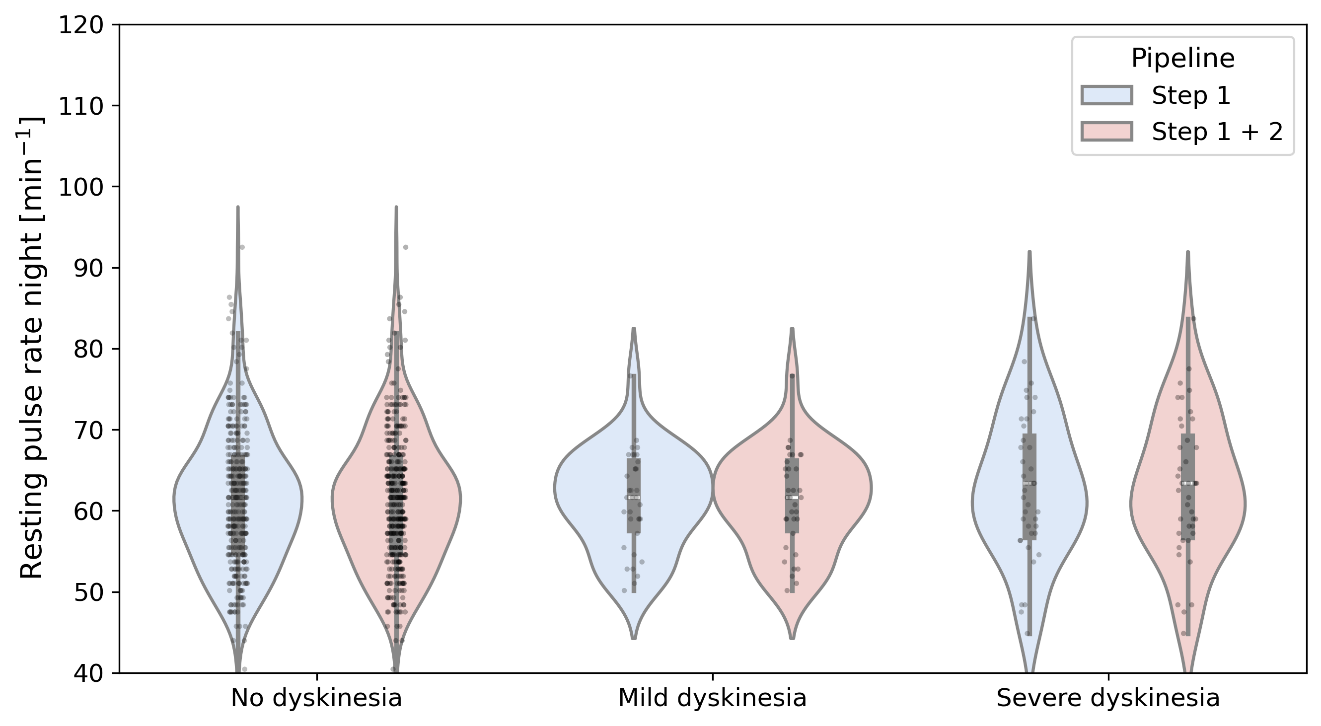

**Supplementary Figure 13: Violins of the resting pulse rate during the night in relation to dyskinesia severity (MDS-UPDRS 4.1 + 4.2) in the first study week. The different colors represent the resting pulse rate when assessing solely PPG morphology (blue) or also incorporating periodic artifact removal using the accelerometer (red). Filtering for periodic motion artifacts does not significantly affect the resting pulse rate in any dyskinesia group.**

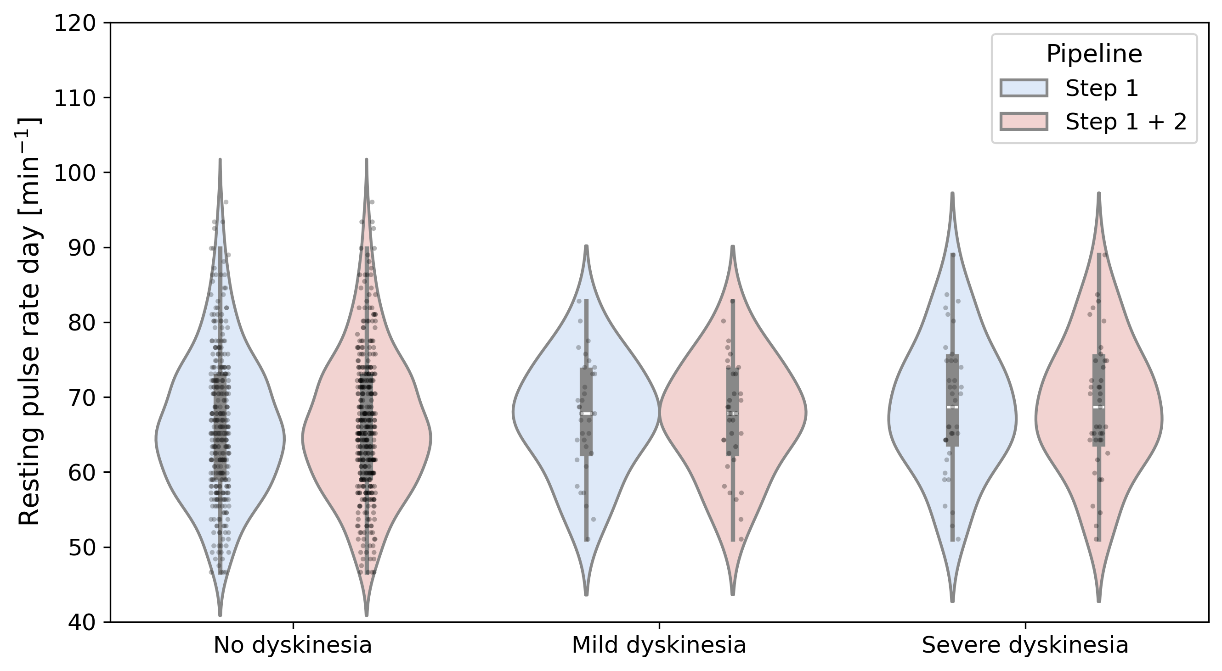

**Supplementary Figure 14: Violins of the resting pulse rate during the day in relation to dyskinesia severity (MDS-UPDRS 4.1 + 4.2) in the first study week. The different colors represent the resting pulse rate when assessing solely PPG morphology (blue) or also incorporating periodic artifact removal using the accelerometer (red). Filtering for periodic motion artifacts does not significantly affect the resting pulse rate in any dyskinesia group.**

#### 2.7 Regression results

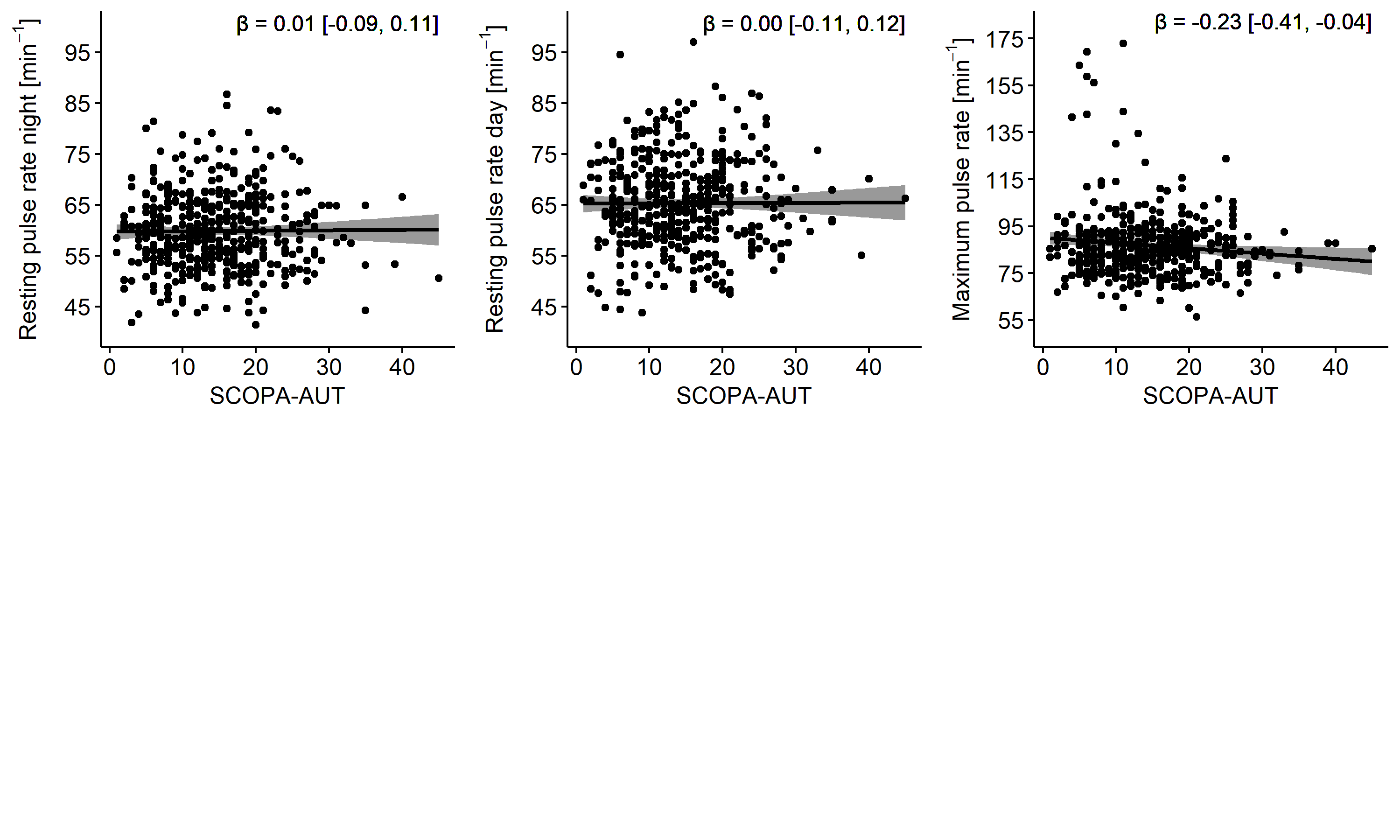

**Supplementary Figure 15. Multivariate regressions between the pulse rate parameters in the second study week and SCOPA-AUT. Every data point is corrected using the following covariates: age, sex, use of betablockers and baseline physical activity.** SCOPA-AUT = SCales for Outcomes in PArkinson’s disease - Autonomic dysfunction.

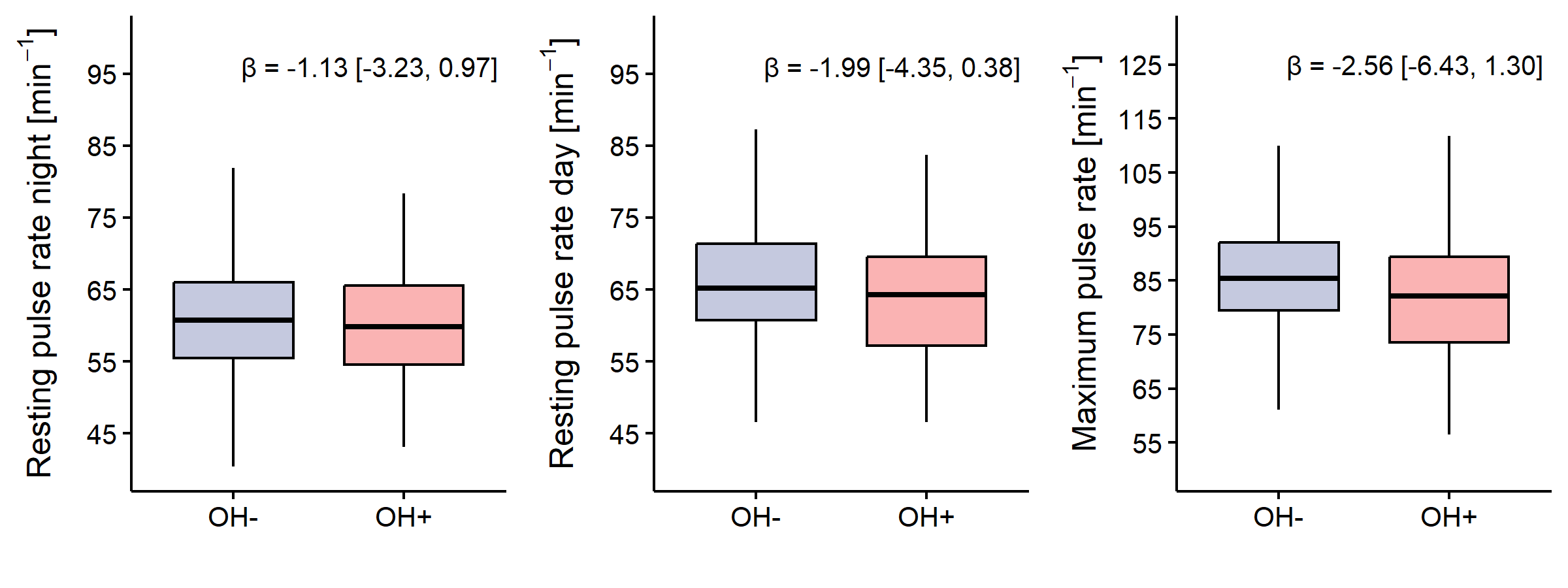
**Supplementary Figure 16. Multivariate regressions between the pulse rate parameters in the first study week and the presence of orthostatic hypotension. Pulse rate parameters are corrected using the following covariates: age, sex, use of betablockers and baseline physical activity.** OH = orthostatic hypotension.

4.	PPG-DaLiA - UCI Machine Learning Repository. https://archive.ics.uci.edu/dataset/495/ppg+dalia.

5.	Scholl, S. Fourier, Gabor, Morlet or Wigner: Comparison of Time-Frequency Transforms.

6.	Wright, P. S. Short-time fourier transforms and Wigner-Ville distributions applied to the calibration of power frequency harmonic analyzers. *IEEE Trans Instrum Meas* **48**, 475–478 (1999).

7.	OToole, J. M. & Boashash, B. Fast and memory-efficient algorithms for computing quadratic time–frequency distributions. *Appl Comput Harmon Anal* **35**, 350–358 (2013).

8.	Charlton, P. H. *et al.* Detecting beats in the photoplethysmogram: benchmarking open-source algorithms. *Physiol Meas* **43**, 085007 (2022).
